# Automated Anatomy-Based Subsegmentation of Pelvic and Proximal Femoral CT: Validation Across Clinically Relevant Regions and Landmarks

**DOI:** 10.64898/2026.05.14.26353237

**Authors:** Mohammed Rashed, Hatem Al-Abdulrahman

**Author notes:** **Corresponding author:** Mohammed Rashed Aly Abdelrahman, **Email:**. **Email addresses of all authors:** Mohammed Rashed, Hatem Alabdulrahman.

## Abstract

**Background:** Automated pelvic CT segmentation has advanced to reliable coarse bone extraction. Yet the structured anatomical hierarchy required for morphometry, fixation planning, bone quality mapping, and arthroplasty workflows remains unachieved. This study developed and validated a fully automated anatomy-informed pipeline that converts standard pelvic CT into a comprehensive, surgeon-readable subsegmentation of the pelvis and proximal femur.

**Methods:** Pelvic CT datasets were retrospectively collected from anonymized archives of hospitals affiliated with the Directorate of Health Affairs, Sharqia, Egypt. After eligibility screening, 757 normal adult cases were processed using a custom one-click 3D Slicer pipeline integrating TotalSegmentator for coarse extraction, followed by deterministic anatomy-based subsegmentation into 81 segments. One hundred randomly selected cases were validated against expert-corrected reference segmentations using Dice similarity coefficient, volume difference, surface distance metrics, and bilateral symmetry analysis.

**Results:** Of 1,316 screened cases, 757 met eligibility criteria. Across 8,100 case-segment observations, the pipeline achieved a mean Dice of 0.9926 ± 0.0465. Complete agreement was observed for the sacrum, ilium, acetabulum, anterior and posterior columns, sciatic buttress, and all landmarks. Relative decreases were confined to boundary-dependent regions. Bilateral symmetry analysis confirmed a median surface agreement of 99.85% within 5 mm.

**Conclusion:** The pipeline demonstrated high accuracy and reproducibility across a large normal adult dataset, establishing a structured anatomical foundation for quantitative pelvic analysis and surgical planning workflows. Clinical feasibility across abnormal anatomy and decision-level applications awaits dedicated validation.

## Introduction

Pelvic CT segmentation is central to trauma assessment, deformity analysis, and preoperative surgical planning in orthopedic practice[1]. It further enables patient-specific extraction of pelvic parameters and optimization of arthroplasty planning and morphological evaluation[2]. However, manual segmentation remains time-consuming and technically dependent on operator expertise[1], [3].

Automated pelvic CT segmentation has been achieved using atlas-based, statistical shape model, and graph-based approaches[1], [4], [5]. These methods primarily produced segmentation of whole pelvic bones or limited hip structures[1], [4], [6], [7]. Deep-learning models have enabled multi-class segmentation of pelvic structures including the sacrum and bilateral hip bones[8], [9].

Recent advances in CT-based pelvic segmentation have produced task-specific outputs such as coarse pelvic bone masks, fracture-fragment masks, and radiotherapy-oriented pelvic bone contours[10], [11], [12], [13], [14]. However, these approaches do not provide a detailed anatomical hierarchy of pelvic and proximal femoral subsegments. Such structured regional mapping could support subregion-specific morphometry, symmetry-based pelvic assessment, bone-quality mapping, fracture localization, fixation-corridor planning, and arthroplasty measurement workflows[15], [16], [17], [18].

The current study therefore developed and validated an automated anatomy-informed CT pipeline that transforms parent pelvic and proximal femoral masks into clinically relevant subregions and landmarks, enabling reproducible mapping and scalable analysis across large normal adult datasets.

## Methods – Patient Selection and CT Acquisition

Pelvic CT datasets were retrospectively collected from fully anonymized imaging archives of multiple hospitals affiliated with the Directorate of Health Affairs in Sharqia, Egypt, under formal authorization for research and healthcare innovation. The datasets were collected between July 2025 and March 2026.

### Inclusion criteria

Adult CT normal scans were included if they encompassed the entire pelvis and proximal femora, extending from at least the L5 vertebral level superiorly to below the lesser trochanters inferiorly. Native CT quality required near-isotropic acquisition, with in-plane voxel size ≤1.0 mm and slice spacing ≤1.25 mm, predefined to preserve reliable 3D reconstruction and segmentation. Cases with marked pelvic malposition on CT were excluded[19]. Malposition was defined as >5° coronal tilt or >5° axial rotation and was applied as a study-defined quality criterion, as pelvic orientation can affect CT-based measurements[20]. Bone quality was additionally screened at L5, where mean trabecular attenuation ≥160 HU was required as an indicator of sufficient bone density for reliable segmentation[21]. Voxel spacing, pelvic positioning, and L5 attenuation were assessed algorithmically using a custom Python-based workflow implemented in 3D Slicer. Morphological abnormalities, fractures, implants, major imaging artifacts, and eligibility decisions were reviewed by an experienced orthopaedic surgeon (Figure 1).

**Figure 1.**
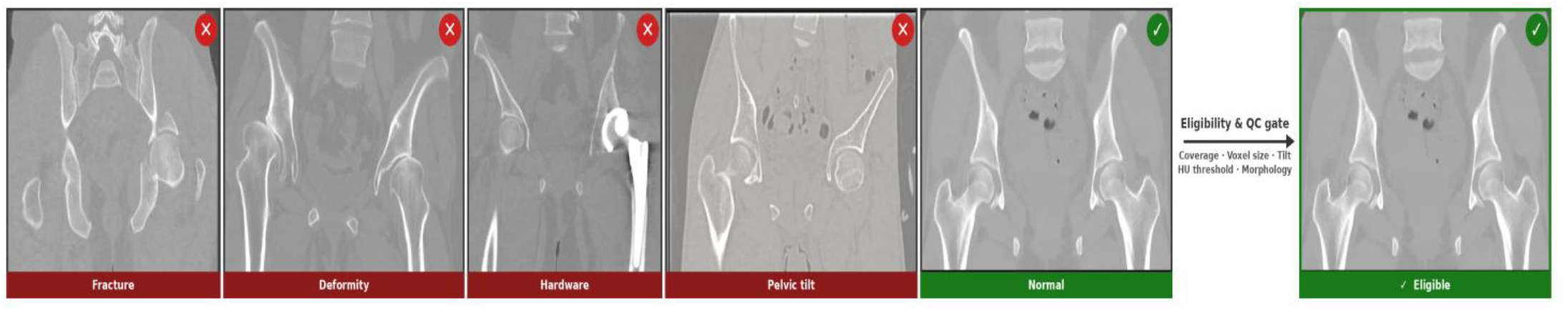
Pelvic CT eligibility and quality-control screening. Representative CT examples show excluded cases and an eligible normal case after assessment of coverage, resolution, bone density, pelvic orientation, and morphology.

**Figure 2.**
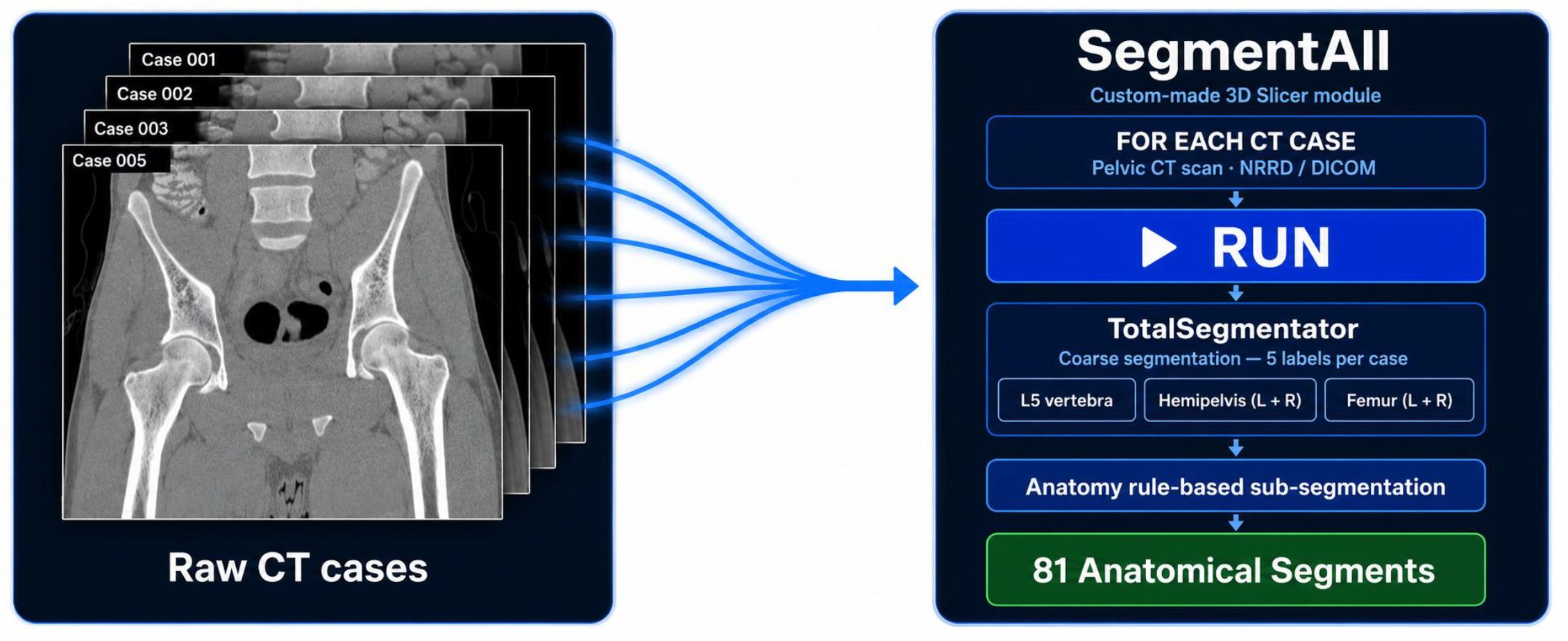
SegmentAll workflow from raw pelvic CTs to the final pelvic and proximal femoral segmentation tree.

### Exclusion criteria

CT scans were excluded if they were obtained in paediatric patients or showed fracture, major deformity, post-surgical change, metallic implants, severe imaging artifacts, incomplete anatomical extent, or failure to satisfy the predefined technical criteria.

### Segmentation workflow

An end-to-end segmentation pipeline for the pelvis and proximal femur was implemented as a custom 3D Slicer module (SegmentAll) [22]. The framework was designed for one-click batch sub segmentation of the full dataset.

TotalSegmentator was integrated to identify the major parent structures required for downstream processing, including L5, the sacrum, both hemipelves, and both femora[9]. The initial sacral mask frequently showed incomplete coverage and was therefore refined using a dedicated rule-based reconstruction step. The confirmed parent masks were then transferred automatically into the rule-based subsegmentation workflow, where deterministic anatomy-based rules were applied across the batch to generate the final subsegmented anatomy (Figure 3). Detailed segmentation logic for individual regions and the anatomical definitions of framework-defined labels are provided in the Supplementary Material.

**Figure 3.**
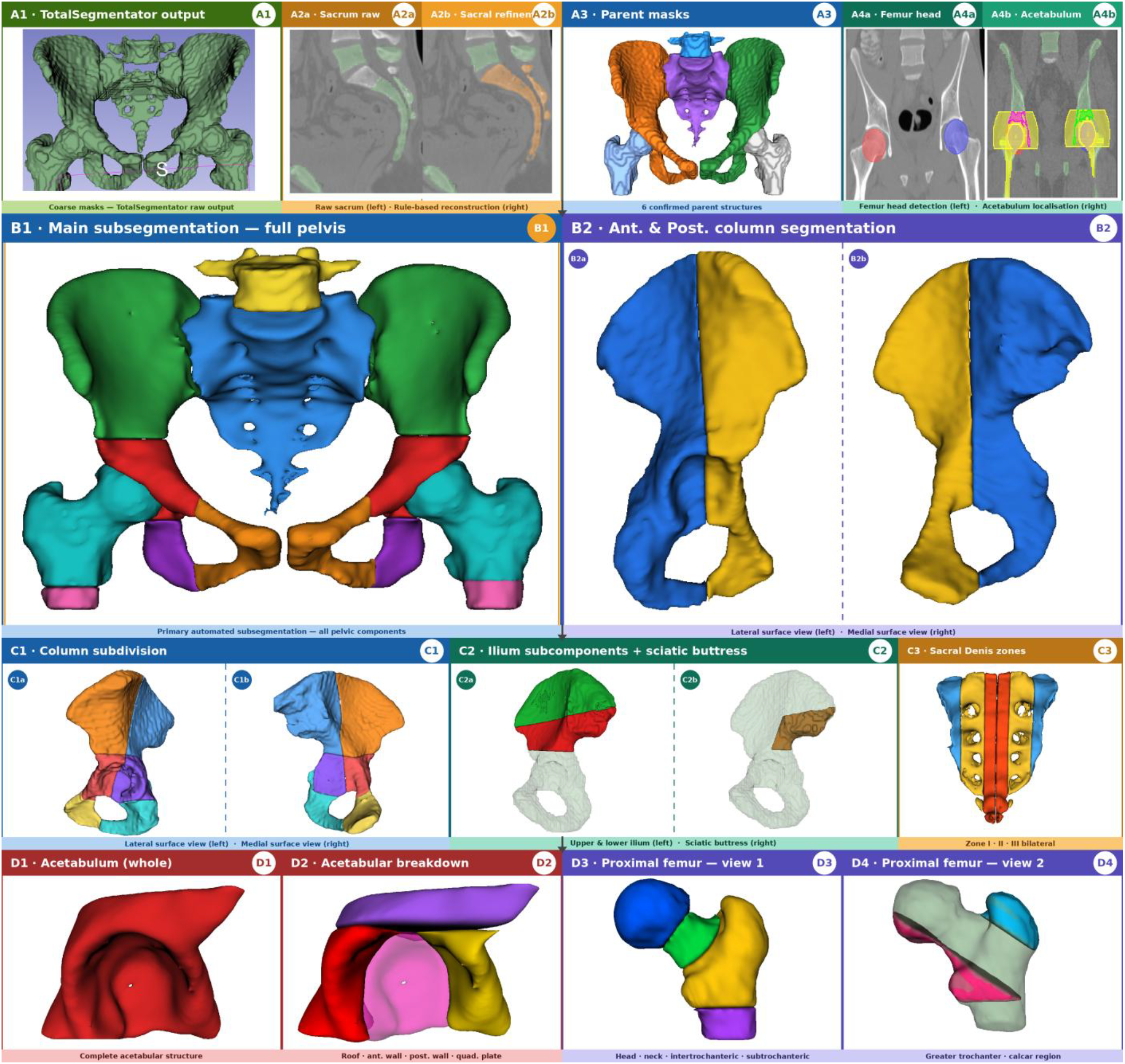
Automated rule-based pelvic CT subsegmentation workflow. (A1) TotalSegmentator coarse masks. (A2) Sacrum raw output (left) and rule-based reconstruction (right). (A3) Six confirmed parent structures. (A4) Bilateral femoral head detection (left) and acetabulum localisation (right) on coronal CT. (B1) Full-pelvis automated subsegmentation result. (B2) Anterior and posterior column segmentation; lateral (left) and medial (right) surface views. (C1) Column subdivision; lateral (left) and medial (right) views. (C2) Ilium subcomponents — upper and lower ilium (left) and sciatic buttress (right). (C3) Sacral Denis zones I–III, bilateral. (D1) Complete acetabulum. (D2) Acetabular breakdown: roof, anterior wall, posterior wall, and quadrilateral plate. (D3) Proximal femur: head, neck, intertrochanteric, and subtrochanteric regions. (D4) Proximal femur: greater trochanter and calcar region.

### Validation

A randomly selected validation subset of 100 eligible cases from the final study cohort was used for detailed accuracy assessment. The automated segmentations were compared against expert-corrected reference segmentations on a per-structure basis. In each case, all segments requiring modification were manually corrected to generate the reference standard. Primary correction was performed by an experienced orthopedic pelvic surgeon, and the final corrected segmentations were subsequently reviewed by a second experienced pelvic surgeon for confirmation. Primary validation included the Dice similarity coefficient (DSC), edited segments (%), absolute percentage volume difference (APVD), center-of-mass distance (CMD), average symmetric surface distance (ASSD), and 95th-percentile Hausdorff distance (HD95). Secondary validation assessed anatomical plausibility by left–right symmetry analysis after contralateral mirroring and rigid surface registration, summarized by root-mean-square surface distance (RMSD) and the proportions of corresponding surface points within 1, 2, 3, 4, and 5 mm.

### Statistical analysis

Exported validation outputs were consolidated and analyzed using a custom external Python script with predefined integrity checks for case number, segment count, primary observations, and secondary symmetry observations. Results were summarized descriptively at the overall, segment, regional, and case levels. Continuous variables were reported as mean ± standard deviation and, where applicable, as median with interquartile range. Right and left observations were pooled within predefined anatomical categories where appropriate. Correction frequency was defined as Dice < 1.0 at the segment level and as the number of corrected segments per case at the case level.

## Results

A total of 1316 pelvic CT cases were screened for eligibility. Of these, 559 cases were excluded: 72 pediatric cases, 218 fracture cases (98 femoral, 69 pelvic-ring, and 51 acetabular fractures), 61 cases with metallic implants or hardware, 32 cases with advanced coxarthrosis or relevant pelvic/hip deformity, and 176 cases with technical or eligibility-related imaging limitations caused by incomplete L5-to-lesser-trochanter coverage of required segmentation components, marked osteoporosis, pelvic malposition/rotation, or inadequate image quality. The final dataset comprised 757 normal adult pelvic CT cases, all processed successfully by the automated segmentation pipeline; 100 cases were randomly selected for detailed primary and secondary validation.

### Overall Primary Segmentation Agreement

The automated segmentation achieved excellent overall agreement with the reference standards. Across 100 validation cases and 8100 analyzed case-segment observations, the mean Dice coefficient was 0.9926 ± 0.0465. Geometric deviations likewise remained low, supporting high overall segmentation accuracy across the validation dataset.

**Table 1.** Overall primary validation metrics across all analyzed segments.

Metrics were calculated across all analyzed case-segment observations in the 100-case validation dataset.
| Metric | Mean ± SD |
| --- | --- |
| Dice | 0.9926 ± 0.0465 |
| VOLdiff (%) | 2.0831 ± 84.2332 |
| COM (mm) | 0.2366 ± 1.9287 |
| ASSD (mm) | 0.0983 ± 1.2200 |
| HD95 (mm) | 0.4299 ± 3.0096 |

### Primary Validation by Major Anatomical and Structural Segments

Performance across the major anatomical and structural segments remained uniformly high. Complete agreement was observed for the sacrum, ilium, acetabulum, anterior column, and posterior column. The lowest values were seen in pubis and ischium, although agreement remained strong overall.

**Table 2.** Primary validation metrics for major anatomical and structural segments.

Right and left measurements were pooled within each anatomical category where applicable; Dice = 1.0000 indicates complete overlap, while VOLdiff, COM, ASSD, and HD95 = 0 indicate no measurable difference. Values are reported as mean ± SD.
| Segment | Dice, mean ± SD | VOLdiff (%), mean ± SD | COM (mm), mean ± SD | ASSD (mm), mean ± SD | HD95 (mm), mean ± SD |
| --- | --- | --- | --- | --- | --- |
| Sacrum | 1.0000 ± 0.0000 | 0.0000 ± 0.0000 | 0.0000 ± 0.0000 | 0.0000 ± 0.0000 | 0.0000 ± 0.0000 |
| Ilium | 1.0000 ± 0.0000 | 0.0000 ± 0.0000 | 0.0000 ± 0.0000 | 0.0000 ± 0.0000 | 0.0000 ± 0.0000 |
| Acetabulum | 1.0000 ± 0.0000 | 0.0000 ± 0.0000 | 0.0000 ± 0.0000 | 0.0000 ± 0.0000 | 0.0000 ± 0.0000 |
| Pubis | 0.9635 ± 0.1214 | 5.2118 ± 15.9714 | 1.4662 ± 7.4970 | 0.5740 ± 4.7014 | 2.2534 ± 11.7066 |
| Ischium | 0.9557 ± 0.0480 | 7.4470 ± 8.2434 | 1.1828 ± 1.4547 | 0.3502 ± 0.4888 | 2.3991 ± 3.8705 |
| Proximal femur | 0.9921 ± 0.0187 | 1.5042 ± 3.5092 | 0.7183 ± 1.7144 | 0.1158 ± 0.2992 | 1.0634 ± 2.8119 |
| Femoral head | 0.9874 ± 0.0273 | 2.1706 ± 4.8190 | 0.4193 ± 0.9333 | 0.1875 ± 0.4055 | 1.3089 ± 2.7701 |
| Anterior column | 1.0000 ± 0.0000 | 0.0000 ± 0.0000 | 0.0000 ± 0.0000 | 0.0000 ± 0.0000 | 0.0000 ± 0.0000 |
| Posterior column | 1.0000 ± 0.0000 | 0.0000 ± 0.0000 | 0.0000 ± 0.0000 | 0.0000 ± 0.0000 | 0.0000 ± 0.0000 |

### Primary Validation by Clinically Relevant Subsegmentation

Clinically relevant subsegments likewise showed high accuracy. The best performance was observed in the sciatic buttress, quadrilateral plate, greater trochanter, and dense sacral compartments, whereas the femoral neck showed the lowest agreement and greatest variability. The remaining subsegments demonstrated strong concordance with the reference standard.

**Table 3.** Primary validation metrics for clinically relevant subsegmentation.

Right and left measurements were pooled within each anatomical category where applicable; Dice = 1.0000 indicates complete overlap, while VOLdiff, COM, ASSD, and HD95 = 0 indicate no measurable difference. Values are reported as mean ± SD.
| Group | Segment | Dice, mean ± SD | VOLdiff (%), mean ± SD | COM (mm), mean ± SD | ASSD (mm), mean ± SD | HD95 (mm), mean ± SD |
| --- | --- | --- | --- | --- | --- | --- |
| Acetabular | Acetabular roof | 0.9944 ± 0.0594 | 0.6998 ± 7.1357 | 0.0643 ± 0.6743 | 0.0386 ± 0.4454 | 0.1237 ± 1.3181 |
| Acetabular | Anterior wall | 0.9946 ± 0.0348 | 1.0312 ± 6.5321 | 0.2144 ± 1.3727 | 0.0387 ± 0.2499 | 0.2354 ± 1.5439 |
| Acetabular | Posterior wall | 0.9958 ± 0.0353 | 0.8034 ± 6.2337 | 0.1109 ± 0.8290 | 0.0368 ± 0.3142 | 0.1935 ± 1.4944 |
| Acetabular | Quadrilateral plate | 0.9996 ± 0.0037 | 0.0925 ± 0.7610 | 0.0394 ± 0.3185 | 0.0055 ± 0.0496 | 0.0292 ± 0.4123 |
| Iliac | Sciatic buttress | 1.0000 ± 0.0000 | 0.0000 ± 0.0000 | 0.0000 ± 0.0000 | 0.0000 ± 0.0000 | 0.0000 ± 0.0000 |
| Femoral | Femoral neck | 0.9276 ± 0.1317 | 8.6454 ± 16.1708 | 1.6022 ± 3.1002 | 1.0297 ± 2.1390 | 4.1879 ± 7.4087 |
| Femoral | Intertrochanteric region | 0.9818 ± 0.0417 | 2.9006 ± 7.8574 | 0.7769 ± 2.1712 | 0.2982 ± 0.9764 | 1.9565 ± 4.7957 |
| Femoral | Calcar region | 0.9713 ± 0.1032 | 3.0364 ± 11.1320 | 1.2338 ± 4.8416 | 0.2444 ± 0.9163 | 0.9428 ± 3.5239 |
| Femoral | Greater trochanter | 0.9985 ± 0.0053 | 0.2857 ± 1.0442 | 0.0621 ± 0.2183 | 0.0251 ± 0.0995 | 0.1204 ± 0.6777 |
| Femoral | Subtrochanteric region | 0.9568 ± 0.1047 | 11.5935 ± 31.6592 | 1.0213 ± 2.4458 | 0.4707 ± 1.1941 | 1.7461 ± 4.1791 |

### Landmark Structures

Landmark segmentation also remained highly accurate. Complete agreement with the reference standard was observed for the symphysis pubis, ASIS, AIIS, PSIS, and iliac crest apex, whereas the lesser trochanter showed slightly lower but still high agreement (0.9883 ± 0.0917).

### Overall Secondary Symmetry Validation

Secondary symmetry analysis confirmed excellent bilateral agreement. The median RMS surface mismatch was 1.2494 mm [1.0456, 1.5772]. Threshold-based agreement increased progressively with wider tolerance levels, reaching a median of 99.8537% within 5 mm.

**Table 4.** Overall secondary symmetry validation metrics.

Secondary symmetry metrics were calculated across all regional bilateral comparisons in the 100-case validation dataset; threshold values indicate the percentage of surface points within each distance tolerance.
| Metric | Mean ± SD | Median [Q1, Q3] |
| --- | --- | --- |
| RMS (mm) | 1.4840 ± 1.5039 | 1.2494 [1.0456, 1.5772] |
| Within 1 mm (%) | 63.4942 ± 12.0345 | 63.0662 [56.0684, 72.4334] |
| Within 2 mm (%) | 88.5457 ± 8.5802 | 90.4710 [85.2930, 94.2608] |
| Within 3 mm (%) | 95.8609 ± 5.8051 | 97.7556 [94.9489, 98.9801] |
| Within 4 mm (%) | 98.0124 ± 4.3905 | 99.3767 [97.7988, 99.8698] |
| Within 5 mm (%) | 98.8264 ± 3.6654 | 99.8537 [99.0414, 99.9933] |

### Secondary Symmetry Validation by Region

Regional symmetry also remained high. The lowest RMS values were observed in the acetabulum, femoral head, and ilium, whereas relatively higher values were seen in the pubis, ischium, and column-based regions.

**Table 5.** Secondary symmetry validation metrics by region.

Regional RMS values summarize bilateral surface agreement for each mirrored anatomical category. Values are reported as mean ± SD and median [Q1, Q3].
| Region | Group | RMS, mean ± SD (mm) | RMS, median [Q1, Q3] (mm) |
| --- | --- | --- | --- |
| Acetabulum | Primary | 1.0307 ± 0.2022 | 1.0014 [0.9043, 1.1029] |
| Femoral head | Primary | 1.0732 ± 0.3138 | 0.9645 [0.8733, 1.2163] |
| Ilium | Primary | 1.2229 ± 0.2298 | 1.1783 [1.0598, 1.3199] |
| Hemipelvis | Primary | 1.3847 ± 0.3013 | 1.3391 [1.2181, 1.4632] |
| Proximal femur | Primary | 1.2992 ± 0.4328 | 1.1863 [1.0846, 1.3337] |
| Ischium | Primary | 1.4377 ± 0.6894 | 1.2701 [1.0029, 1.7670] |
| Pubis | Primary | 2.0513 ± 4.1376 | 1.2833 [0.8760, 1.9214] |
| Anterior column | Secondary | 1.8778 ± 0.6937 | 1.6734 [1.4014, 2.1497] |
| Posterior column | Secondary | 1.9781 ± 0.8773 | 1.6985 [1.4349, 2.2086] |

### Segment-Level Correction Frequency

Manual correction was concentrated in a limited subset of segments. The ischium and femoral neck required correction most often and were associated with the lowest mean Dice values among frequently edited regions. The intertrochanteric region, femoral head, proximal femur, and subtrochanteric region also required correction relatively often, whereas most remaining segments required only occasional or isolated editing.

**Table 6.**
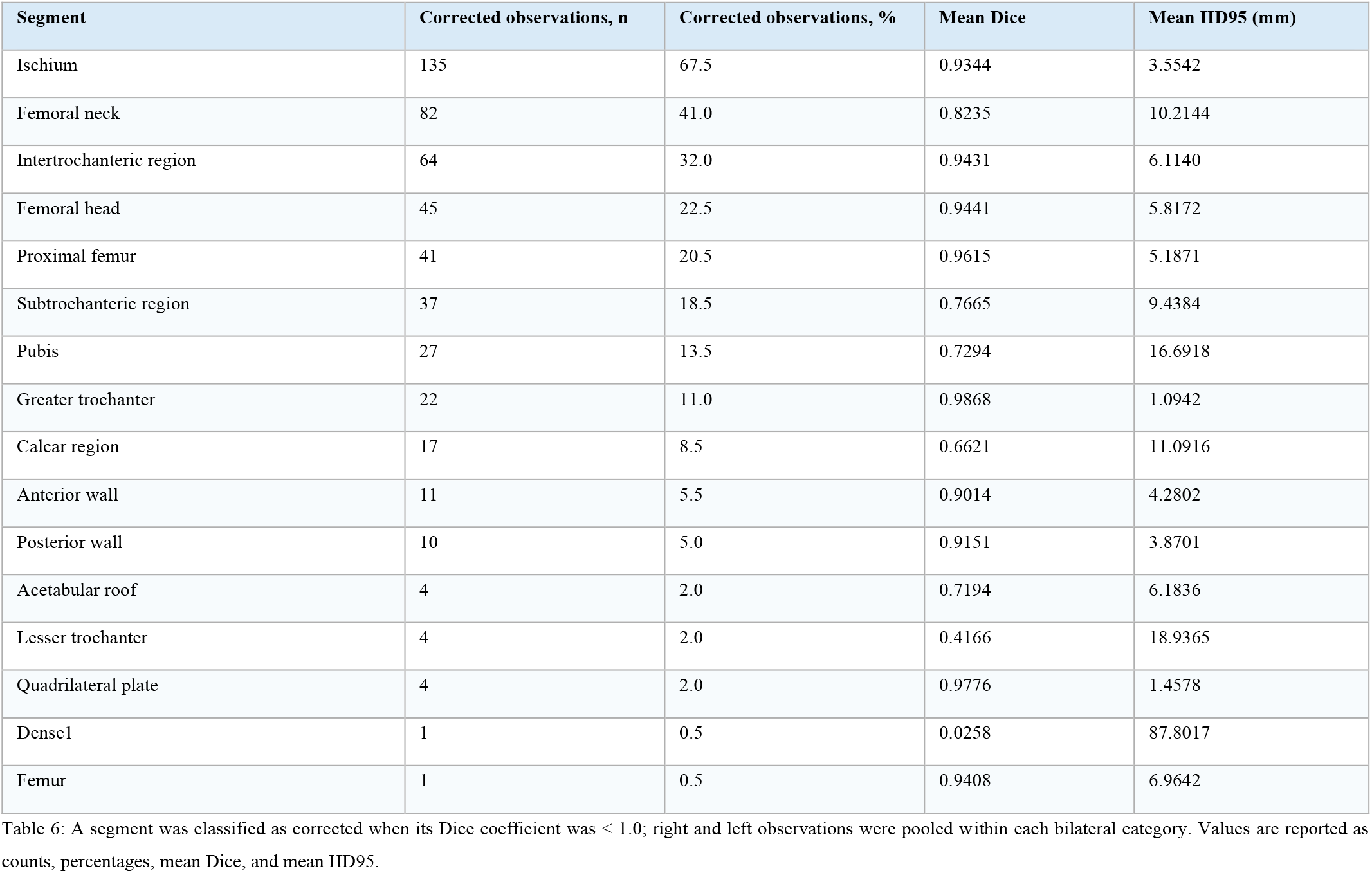
Segment-level correction frequency across the 100-case validation dataset.

### Case-Level Correction Burden

At the case level, the overall editing burden remained limited. The median number of corrected segments per case was 4.5, with a maximum of 19 corrected segments in any individual case. Four cases required no correction, and most cases required correction in only a small number of segments.

**Table 7.**
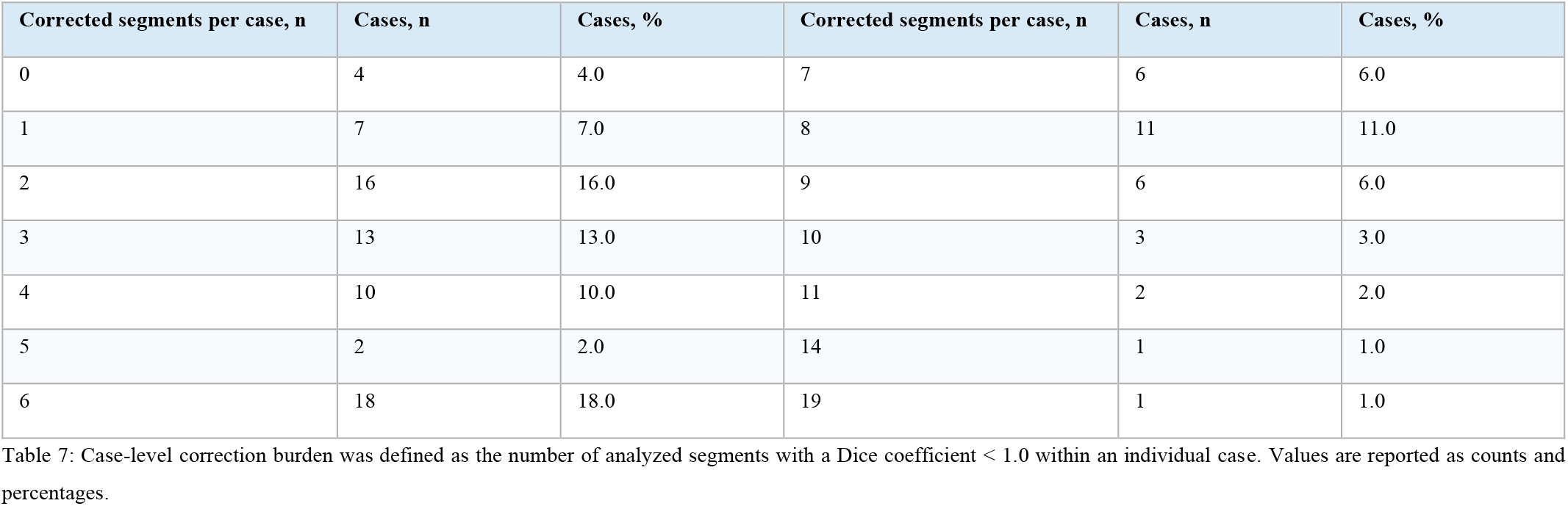
Case-level correction burden distribution across the 100-case validation dataset.

Case-level correction burden was defined as the number of analyzed segments with a Dice coefficient $< 1.0$ within an individual case. Values are reported as counts and percentages.
| Corrected segments per case, n | Cases, n | Cases, % | Corrected segments per case, n | Cases, n | Cases, % |
| --- | --- | --- | --- | --- | --- |
| 0 | 4 | 4.0 | 7 | 6 | 6.0 |
| 1 | 7 | 7.0 | 8 | 11 | 11.0 |
| 2 | 16 | 16.0 | 9 | 6 | 6.0 |
| 3 | 13 | 13.0 | 10 | 3 | 3.0 |
| 4 | 10 | 10.0 | 11 | 2 | 2.0 |
| 5 | 2 | 2.0 | 14 | 1 | 1.0 |
| 6 | 18 | 18.0 | 19 | 1 | 1.0 |

## Discussion

Pelvic anatomy poses a distinct three-dimensional analytical challenge [23]. CT remains the gold standard for detailed evaluation of the pelvis [24]. Its assessment requires integration of diverse findings, including acetabular measurements, pelvic alignment, bone stock, symmetry, and fracture classification [20], [25], [26]. Therefore, segmentation is particularly valuable in pelvic CT, as it converts complex anatomy into structures that can be consistently visualized, localized, and measured [27].

Manual workflows remain a major barrier in pelvic CT analysis[3], [28]. Previous reports showed that CT-based 3D model segmentation took 90–120 minutes for each hip joint, while manual CT-based THA planning averaged 185.40 ± 21.76 minutes per case [3], [28]. Additionally, prior data showed that some pelvic landmarks were frequently mislabeled by 10 mm or more [25]. CT-based pelvic analysis is also challenging in several clinically important and research-oriented tasks [2], [29]. These include pelvimetric extraction, fracture classification, fixation planning, deformity correction, and large-scale generation of structured anatomical data for research use [2], [30]. Together, these limitations underscore the value of automated segmentation for structured and reproducible pelvic CT analysis [27].

Earlier pelvic CT studies established the feasibility of atlas-based segmentation, landmark transfer, and targeted hip-joint extraction [1], [5]. [6]. [4] [31] [7]. Recent CT-based pelvic segmentation work has remained task-specific. Coarse bone-segmentation approaches produced lumbar spine, sacrum, bilateral hemipelves, femoral, or whole-pelvis labels.[8], [9], [11], [12], [32].Trauma-focused pipelines targeted pelvic fracture masks, fragment segmentation, or fracture-reduction planning[10], [14], [33]. Measurement- and landmark-based frameworks automated selected outputs such as 3D pelvimetry, acetabular lunate-surface analysis, or landmark detection in defective pelvis CTs[2], [34], [35].Therefore, current literature supports automated pelvic CT analysis for defined tasks, but a unified fully automated sub segmentation framework of normal pelvic and proximal femoral subregions remains insufficiently established.

In the current study, sub segmentation was reframed from coarse bone-mask extraction into an orthopaedic anatomical mapping system. Each output region was selected and defined according to anatomical units with direct clinical applicability in pelvic assessment and surgical planning. The acetabulum is the osseous convergence of the ilium, ischium, and pubis—the three fused components of the hemipelvis[29]. Accordingly, these four structures were defined as the principal pelvic components and then organized into anterior and posterior column divisions, reflecting the structural logic of pelvic surgical anatomy[36]. The hierarchy was then extended to finer surgically relevant regions, for instance, the acetabular walls and quadrilateral plate support assessment of acetabular containment and column integrity[37], [38]. The sciatic buttress and femoral calcar provide pelvic and proximal femoral reference regions for hip load transfer and regional bone stock [39], [40], [41]. Denis-based sacral zones convert the sacrum from a single mask into a fracture-oriented map of alar, foraminal, and central involvement[42]. This segmentation design is particularly relevant for future fracture mapping, regional morphometry, and planning-oriented CT analysis, where interpretation depends on precise anatomical location rather than whole-bone volume alone.

Pelvic CT segmentation remains boundary-sensitive even before fine sub segmentation. Recent work has described challenges related to small targets, thin boundaries, and separation of adjacent hip bone segments [10]. In the present study, high agreement was maintained across a dense set of rule-defined anatomical subdivisions. Agreement was strongest in stable reference regions, including the sacrum, ilium, acetabulum, and anterior and posterior columns. As the workflow moved toward finer regional subdivision, performance remained stable in representative acetabular, iliac, sacral, and proximal-femoral subregions, such as the acetabular walls, sciatic buttress, dense sacral compartments, femoral head, and greater trochanter. Relative decreases were limited to boundary-dependent or bridging regions, mainly the femoral neck, ischium, pubis, subtrochanteric region, calcar region, and intertrochanteric region, where anatomical continuity makes discrete voxel-level separation less straightforward. This boundary profile explains why correction frequency was concentrated in a small subset of transition-related segments rather than distributed uniformly across the anatomical tree.

Landmark segmentation showed similarly high agreement and added practical value beyond surface accuracy. Stable landmarks such as the ASIS, AIIS, PSIS, iliac crest apex, symphysis pubis, and lesser trochanter provide fixed reference points for pelvic orientation, coordinate construction, bilateral comparison, angular measurement, and planning outputs. Thus, the validation results indicate that the pipeline is not only accurate at the mask level, but also reliable as a structured anatomical reference system for downstream pelvic CT analysis.

Symmetry analysis served as a secondary plausibility check for segmentation consistency. In the present study, contralateral mirroring and rigid surface registration were applied across 100 CT cases and paired regions including the acetabulum, femoral head, ilium, innominate, ischium, proximal femur, pubis, and anterior and posterior columns; median surface agreement reached 89.9438% within 2 mm and 99.8108% within 5 mm. These findings are consistent with previous 3D studies of intact pelvic symmetry[43], [44], [45]. Ead et al. analyzed 14 pelvic CT scans using Mimics-based 3D reconstruction and Geomagic deviation analysis, reporting an RMS deviation of 1.14 ± 0.26 mm and 91.9 ± 5.55% of surface points within ±2 mm[43]. Zheng et al. analyzed 50 pelvic CT scans using Mimics, Geomagic, and closest-point alignment, reporting a mean surface deviation of 1.15 ± 0.16 mm and 90.82 ± 4.67% of points within the permissible ±2 mm range[44]. Bakhshayesh et al. analyzed ten healthy adult CT scans using STL reconstruction, hemipelvis mirroring, volume fusion, and CT motion analysis, showing that right–left differences remained within approximately 3 mm translation and 2° rotation[45]. These studies establish bilateral pelvic symmetry as a reliable reference concept; the current study extends this concept from whole-pelvis comparison to region-level segmentation, supporting its use for side-to-side quality control, normal-variation assessment, and detection of abnormal asymmetry.

Beyond validation, the segmentation framework was integrated into a custom one-click 3D Slicer workflow to assess its feasibility for quantitative analysis and clinically relevant secondary applications. These included automated regional bone-quality assessment in predefined load-bearing areas, selected pelvic and proximal femoral morphometric measurements, and planning-oriented workflows for fracture fixation and arthroplasty. The same framework may also support structured annotation of abnormal or excluded cases, where automated segmentation followed by selective manual refinement could accelerate the creation of curated datasets for future deep-learning models. These outputs remain extensions of the core pipeline and represent planned directions for follow-up studies; each requires application-specific validation before clinical use. Technical details for these secondary applications are provided in the Supplementary Material..

## Strengths and limitations

To our knowledge, this is the first automated pelvic CT framework to move beyond coarse bone segmentation or single-task measurement and provide an expert-validated, anatomy-based orthopedic map of the pelvis and proximal femur. Its main strength is that pelvic CT segmentation is treated as a reproducible anatomical framework, preserving clinically meaningful regions, landmarks, and spatial relationships rather than generating isolated masks. This design provides a transparent and reusable foundation for downstream morphometric, measurement, and planning workflows. The main limitation is that the pipeline is deterministic and was validated in normal adult CT scans; therefore, performance in fractures, deformity, severe degeneration, hardware, or marked unilateral morphometric alteration remains unproven and requires dedicated evaluation before abnormal-cohort or decision-level clinical use.

## Conclusion

This study validates an automated pelvic CT framework that turns normal pelvic and proximal femoral anatomy into a structured, surgeon-readable computational map. Its key advance is the automated preservation of clinically relevant regions, landmarks, and spatial relationships that are usually lost when pelvic CT is reduced to coarse bone masks. By converting segmentation into an anatomical layer, the framework creates a reusable foundation for quantitative analysis, planning-oriented workflows, and future pathology-aware dataset generation. Further validation is required before extending this framework to abnormal anatomy or clinical decision making.

## Supporting information

Supplementary Clinical Applications and Feasibility Analysis.

## Data Availability

The anonymized imaging data analyzed in this study are not publicly available due to institutional data-governance restrictions, but derived study data and analysis outputs are available from the corresponding author upon reasonable request and subject to institutional approval.

## Abbreviation list

AIIS: Anterior inferior iliac spine
APVD: Absolute percentage volume difference
ASSD: Average symmetric surface distance
ASIS: Anterior superior iliac spine
CMD: Center-of-mass distance
DSC: Dice similarity coefficient
HD95: 95th-percentile Hausdorff distance
HU: Hounsfield unit
PSIS: Posterior superior iliac spine
RMSD: Root-mean-square surface distance
RMS: Root-mean-square
THA: Total hip arthroplasty
VOLDiff: Absolute volume difference (%)

## Statements and Declarations

### Ethics Approval and Consent to Participate

All methods were carried out in accordance with relevant guidelines and regulations. This study was performed in line with the principles of the Declaration of Helsinki. Data were collected retrospectively from fully anonymized imaging archives under formal authorization for research and healthcare innovation. Due to the retrospective nature of the study and the use of fully anonymized data, the requirement for written informed consent was waived by the ethics committee.

### Consent to Publish

Not applicable.

## Authors’ Contributions

### Mohammed Rashed

Conceptualization, study design, data collection, eligibility screening, pipeline development, framework design, segmentation workflow implementation, validation analysis, and manuscript writing.

### Hatem Al-Abdulrahman

Data evaluation, secondary observer agreement assessment, and review of the reference-standard segmentations.

## Availability of Data and Materials

The datasets collected during this study will be stored securely at our institution for a period of 10 years, after which they will be deleted in accordance with institutional data retention policy. All datasets analyzed or generated during this study are available from the corresponding author upon reasonable request.

## Acknowledgements

The authors are not compensated and there are no institutional subsidies, corporate affiliations, or funding sources supporting this work. The authors report no conflicts of interest concerning the materials, methods, or findings described in this article. The work described has not been published previously, is not under consideration for publication elsewhere, and its submission has been approved by all co-authors.

## References

[1] H. Seim, D. Kainmueller, M. Heller, H. Lamecker, S. Zachow, and H.-C. Hege, “Automatic Segmentation of the Pelvic Bones from CT Data Based on a Statistical Shape Model.”

[2] J. Shao, Q. Wu, Y. Zhang, C. Liu, X. Huo, and C. Wang, “Automatic 3D pelvimetry framework in CT images and its validation,” Sci. Rep., vol. 14, no. 1, Dec. 2024, doi: 10.1038/s41598-024-72123-6.

[3] Y. Cheng, S. Zhou, Y. Wang, C. Guo, J. Bai, and S. Tamura, “Automatic segmentation technique for acetabulum and femoral head in CT images,” Pattern Recognit., vol. 46, no. 11, pp. 2969–2984, Nov. 2013, doi: 10.1016/j.patcog.2013.04.006.

[4] “Atlas-based Recognition of Anatomical Structures and Landmarks and the Automatic Computation of Orthopedic Parameters,” 2004.

[5] R. A. Zoroofi et al., “Automated Segmentation of Acetabulum and Femoral Head from 3-D CT Images,” IEEE Transactions on Information Technology in Biomedicine, vol. 7, no. 4, pp. 329–343, Dec. 2003, doi: 10.1109/TITB.2003.813791.

[6] C. Chu, J. Bai, X. Wu, and G. Zheng, “MASCG: Multi-Atlas Segmentation Constrained Graph method for accurate segmentation of hip CT images,” Med. Image Anal., vol. 26, no. 1, pp. 173–184, Dec. 2015, doi: 10.1016/j.media.2015.08.011.

[7] P. Liu et al., “Deep learning to segment pelvic bones: large-scale CT datasets and baseline models,” Int. J. Comput. Assist. Radiol. Surg., vol. 16, no. 5, pp. 749–756, May 2021, doi: 10.1007/s11548-021-02363-8.

[8] J. Wasserthal et al., “TotalSegmentator: Robust Segmentation of 104 Anatomic Structures in CT Images • Content codes,” Radiol. Artif. Intell., vol. 5, no. 5, 2023, doi: 10.5281/zenodo.6802613.

[9] T. Qin, X. Yuan, X. Yan, L. Xu, and Y. Liu, “HGSF-NET: AUTOMATIC PELVIC FRACTURE CT SEGMENTATION WITH 2.5D NEIGHBORHOOD CONTEXT AND ADAPTIVE DETAIL GUIDANCE,” J. Mech. Med. Biol., Apr. 2026, doi: 10.1142/S0219519426400282.

[10] S. J. Nesheli et al., “Transformer-based and CNN-based models for clinically effective 2D and 3D pelvic bone segmentation in CT imaging,” BMC Musculoskelet. Disord., Dec. 2025, doi: 10.1186/s12891-025-09444-8.

[11] C. Li, L. Chen, Q. Liu, and J. Teng, “Lumbar and pelvic CT image segmentation based on cross-scale feature fusion and linear self-attention mechanism,” Sci. Rep., vol. 15, no. 1, Dec. 2025, doi: 10.1038/s41598-025-13569-0.

[12] K. Kosaka et al., “Automatic pelvic bone segmentation for radiotherapy using diagnostic imaging workstation,” Reports of Practical Oncology and Radiotherapy, Feb. 2025, doi: 10.5603/rpor.109519.

[13] Y. Liu et al., “Automatic pelvic fracture segmentation: a deep learning approach and benchmark dataset,” Front. Med. (Lausanne)., vol. 12, 2025, doi: 10.3389/fmed.2025.1511487.

[14] “An_End-to-End_Geometry-Based_Pipeline_for_Automatic_Preoperative_Surgical_Planning_of_Pelvic_Fracture_Reduction_and_Fixation”.

[15] N. Ramadanov and S. Zabler, “Ramadanov–Zabler Safe Zone for Sacroiliac Screw Placement: A CT-Based Computational Pilot Study,” J. Clin. Med., vol. 14, no. 10, May 2025, doi: 10.3390/jcm14103567.

[16] T. J. Yang and W. Qian, “AI-Assisted 3D Planning of CT Parameters for Personalized Femoral Prosthesis Selection in Total Hip Arthroplasty,” Ther. Clin. Risk Manag., vol. 21, pp. 905–916, 2025, doi: 10.2147/TCRM.S521755.

[17] T. J. Yang et al., “CT Hounsfield units in assessing bone and soft tissue quality in the proximal femur: A systematic review focusing on osteonecrosis and total hip arthroplasty,” PLoS One, vol. 20, no. 3 March, Mar. 2025, doi: 10.1371/journal.pone.0319907.

[18] X. B. Wu et al., “Printed three-dimensional anatomic templates for virtual preoperative planning before reconstruction of old pelvic injuries: Initial results,” Chin. Med. J. (Engl)., vol. 128, no. 4, pp. 477–482, 2015, doi: 10.4103/0366-6999.151088.

[19] H. J. P. van Bosse, D. Lee, E. R. Henderson, D. A. Sala, and D. S. Feldman, “Pelvic Positioning Creates Error in CT Acetabular Measurements,” Clin. Orthop. Relat. Res., vol. 469, no. 6, 2011, [Online]. Available: https://journals.lww.com/clinorthop/fulltext/2011/06000/pelvic_positioning_creates_error_in_ct_acetabular.23.aspx

[20] P. J. Pickhardt, B. D. Pooler, T. Lauder, A. M. del Rio, R. J. Bruce, and N. Binkley, “Opportunistic Screening for Osteoporosis Using Abdominal Computed Tomography Scans Obtained for Other Indications,” Ann. Intern. Med., vol. 158, no. 8, pp. 588–595, Apr. 2013, doi: 10.7326/0003-4819-158-8-201304160-00003.

[21] A. Fedorov et al., “3D Slicer as an image computing platform for the Quantitative Imaging Network,” Magn. Reson. Imaging, vol. 30, no. 9, pp. 1323–1341, Nov. 2012, doi: 10.1016/j.mri.2012.05.001.

[22] C. Arand et al., “3D statistical model of the pelvic ring – a CT-based statistical evaluation of anatomical variation,” J. Anat., vol. 234, no. 3, pp. 376–383, Mar. 2019, doi: 10.1111/joa.12928.

[23] E. Leone et al., “Imaging Review of Pelvic Ring Fractures and Its Complications in High-Energy Trauma,” Feb. 01, 2022, Multidisciplinary Digital Publishing Institute (MDPI). doi: 10.3390/diagnostics12020384.

[24] J. Hêches et al., “Accuracy and Reliability of Pelvimetry Measures Obtained by Manual or Automatic Labeling of Three-Dimensional Pelvic Models,” J. Clin. Med., vol. 13, no. 3, Feb. 2024, doi: 10.3390/jcm13030689.

[25] A. Gänsslen, J. Tonetti, and T. Pohlemann, “Algorithms in acetabular fracture classifications,” Arch. Orthop. Trauma Surg., vol. 144, no. 10, pp. 4655–4665, Oct. 2024, doi: 10.1007/s00402-024-05599-6.

[26] H. Yu, H. Wang, Y. Shi, K. Xu, X. Yu, and Y. Cao, “The segmentation of bones in pelvic CT images based on extraction of key frames,” BMC Med. Imaging, vol. 18, no. 1, May 2018, doi: 10.1186/s12880-018-0260-x.

[27] G. Zeng et al., “MRI-based 3D models of the hip joint enables radiation-free computer-assisted planning of periacetabular osteotomy for treatment of hip dysplasia using deep learning for automatic segmentation,” Eur. J. Radiol. Open, vol. 8, Jan. 2021, doi: 10.1016/j.ejro.2020.100303.

[28] X. Chen et al., “Development and Validation of an Artificial Intelligence Preoperative Planning System for Total Hip Arthroplasty,” Front. Med. (Lausanne)., vol. 9, Mar. 2022, doi: 10.3389/fmed.2022.841202.

[29] D. A. Lawrence, K. Menn, M. Baumgaertner, and A. H. Haims, “Acetabular fractures: Anatomic and clinical considerations,” Sep. 2013. doi: 10.2214/AJR.12.10470.

[30] J. Zwingmann, G. Konrad, E. Kotter, N. P. Südkamp, and M. Oberst, “Computer-navigated Iliosacral Screw Insertion Reduces Malposition Rate and Radiation Exposure,” Clin. Orthop. Relat. Res., vol. 467, no. 7, 2009, [Online]. Available: https://journals.lww.com/clinorthop/fulltext/2009/07000/computer_navigated_iliosacral_screw_insertion.27.aspx

[31] C. Chu, C. Chen, L. Liu, and G. Zheng, “FACTS: Fully Automatic CT Segmentation of a Hip Joint,” Ann. Biomed. Eng., vol. 43, no. 5, pp. 1247–1259, May 2015, doi: 10.1007/s10439-014-1176-4.

[32] A. Diaz-Pinto et al., “MONAI Label: A framework for AI-assisted interactive labeling of 3D medical images,” Med. Image Anal., vol. 95, Jul. 2024, doi: 10.1016/j.media.2024.103207.

[33] Y. Liu et al., “Preoperative fracture reduction planning for image-guided pelvic trauma surgery: A comprehensive pipeline with learning,” Med. Image Anal., vol. 102, May 2025, doi: 10.1016/j.media.2025.103506.

[34] X. Zhao et al., “Defect-adaptive landmark detection in pelvis CT images via personalized structure-aware learning,” Computerized Medical Imaging and Graphics, vol. 127, Jan. 2026, doi: 10.1016/j.compmedimag.2025.102693.

[35] S. De Raedt et al., “Lunate extract: fully automatic acetabular lunate segmentation and hip angle measurements,” Acta radiol., vol. 66, no. 11, pp. 1208–1216, Nov. 2025, doi: 10.1177/02841851251359649.

[36] T. B. Alton and A. O. Gee, “Classifications in brief: Letournel classification for acetabular fractures,” Clin. Orthop. Relat. Res., vol. 472, no. 1, pp. 35–38, 2014, doi: 10.1007/s11999-013-3375-y.

[37] “scheinfeld-et-al-2015-acetabular-fractures-what-radiologists-should-know-and-how-3d-ct-can-aid-classification”.

[38] G. White, N. K. Kanakaris, O. Faour, J. A. Valverde, M. A. Martin, and P. V. Giannoudis, “Quadrilateral plate fractures of the acetabulum: An update,” Feb. 2013. doi: 10.1016/j.injury.2012.10.010.

[39] D. Dreizin, C. A. LeBedis, and J. W. Nascone, “Imaging Acetabular Fractures,” Radiologic Clinics, vol. 57, no. 4, pp. 823–841, Jul. 2019, doi: 10.1016/j.rcl.2019.02.004.

[40] Q. Zhang et al., “The role of the calcar femorale in stress distribution in the proximal femur.,” Orthop. Surg., vol. 1, no. 4, pp. 311–316, 2009, doi: 10.1111/j.1757-7861.2009.00053.x.

[41] R. G. Stiles, C. J. Laverina, D. Resnick, and F. R. Convery, “The Calcar Femorale An Anatomic, Radiologic, and Surgical Correlative Study,” Invest. Radiol., vol. 25, no. 12, 1990, [Online]. Available: https://journals.lww.com/investigativeradiology/fulltext/1990/12000/the_calcar_femorale_an_anatomic,_radiologic,_and.7.aspx

[42] J. M. Rizkalla, T. Lines, and S. Nimmons, “Classifications in Brief: The Denis Classification of Sacral Fractures,” Clin. Orthop. Relat. Res., vol. 477, no. 9, 2019, [Online]. Available: https://journals.lww.com/clinorthop/fulltext/2019/09000/classifications_in_briefthe_denis_classification.36.aspx

[43] M. S. Ead, L. Westover, S. Polege, S. McClelland, J. L. Jaremko, and K. K. Duke, “Virtual reconstruction of unilateral pelvic fractures by using pelvic symmetry,” Int. J. Comput. Assist. Radiol. Surg., vol. 15, no. 8, pp. 1267–1277, Aug. 2020, doi: 10.1007/s11548-020-02140-z.

[44] Q. Zheng et al., “Investigation of pelvic symmetry: A systematic analysis using computer aided design software,” Health Care Science, vol. 2, no. 1, pp. 36–44, Feb. 2023, doi: 10.1002/hcs2.25.

[45] P. Bakhshayesh, A. Zaghloul, B. M. Sephton, and A. Enocson, “A novel 3D technique to assess symmetry of hemi pelvises,” Sci. Rep., vol. 10, no. 1, Dec. 2020, doi: 10.1038/s41598-020-75884-y.

