## Supplementary Clinical Applications and Feasibility Analysis. for "Automated Anatomy-Based Subsegmentation of Pelvic and Proximal Femoral CT: Validation Across Clinically Relevant Regions and Landmarks"

### Clinical and research applications of detailed pelvic CT subsegmentation

Detailed pelvic CT subsegmentation transforms pelvic anatomy from a visual output into a quantitative substrate for measurement, planning, and dataset generation. The same validated segmentation output was reused to derive coverage maps, morphometric indices, bone-quality estimates, arthroplasty-planning parameters, and deep-learning labels. These applications demonstrate feasibility only, and each requires dedicated validation before clinical decision use.

#### Automated pelvimetric extraction

The finalized segmentation tree was used to extract standardized pelvic and proximal femoral measurements directly from anatomical regions and landmarks.

#### Acetabular and femoral-head coverage

Acetabular coverage describes the three-dimensional containment of the femoral head by the acetabulum. [1]

- **3D coverage mapping**

Three-dimensional acetabular and femoral-head coverage can be quantified as surface-area containment and reported globally, regionally by quadrant, and radially by clockface position. [1] Recent CT-based dysplasia work also emphasizes the value of quantifying femoral-head coverage as a three-dimensional surface-area measure [2]. Manual 3D coverage analysis remains difficult to standardize because reproducible assessment requires segmentation, pelvic alignment, acetabular surface definition, regional localization, and comparison with reference values [3].

In the present workflow, acetabular and femoral-head coverage were derived from the finalized Pelvis Segmentation Tree by analyzing the spatial relationship between the segmented acetabular and femoral-head surfaces in a fixed pelvic orientation. The acetabular-coverage output estimates the acetabular surface region participating in femoral-head containment. The femoral-head coverage output estimates the covered femoral-head surface globally and within predefined regional sectors. Quadrant percentages and clockface localization identify whether coverage abnormality is anterior, posterior, lateral, or combined, converting a single projection-dependent angle into a directional 3D containment map[1] (Figure 1).

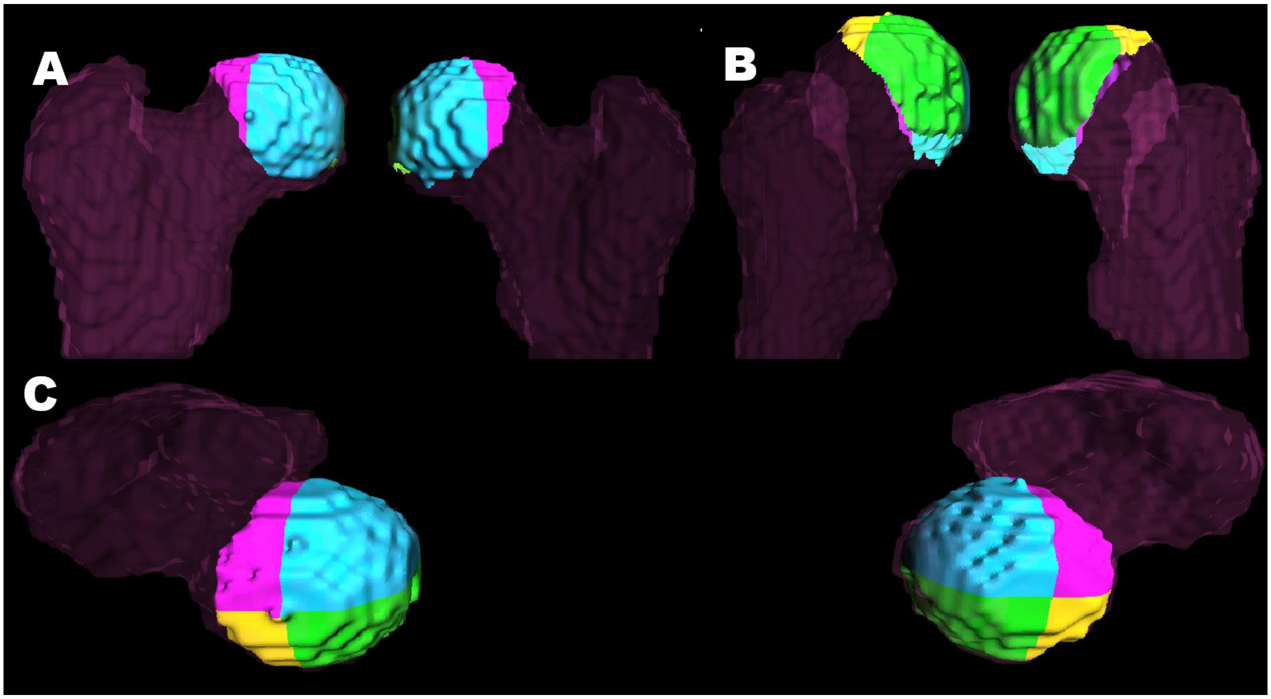


**Figure 1. Surface-based acetabular and femoral-head coverage maps with quadrant-percentage and clockface output.** A 3D frontal view; B, posterior view; C, superior view.

- **Coverage-related angular parameters**

Coverage-related angular parameters were added to preserve familiar clinical interpretation beside the surface maps. Lateral center edge angel (LCEA) quantifies superolateral femoral-head coverage, and recent CT-comparison studies show that radiographs may underestimate LCEA compared with 3D CT [4]. In the present workflow, **LCEA** was derived from the segmented acetabulum and femoral head by extracting the closest acetabular patch, fitting a 3D circle from lateral, inferior, anterior, and posterior patch points, defining the 12-o'clock point from the circle plane, and calculating the angle to the femoral-head center in a fixed AP/coronal plane (Figure 2).

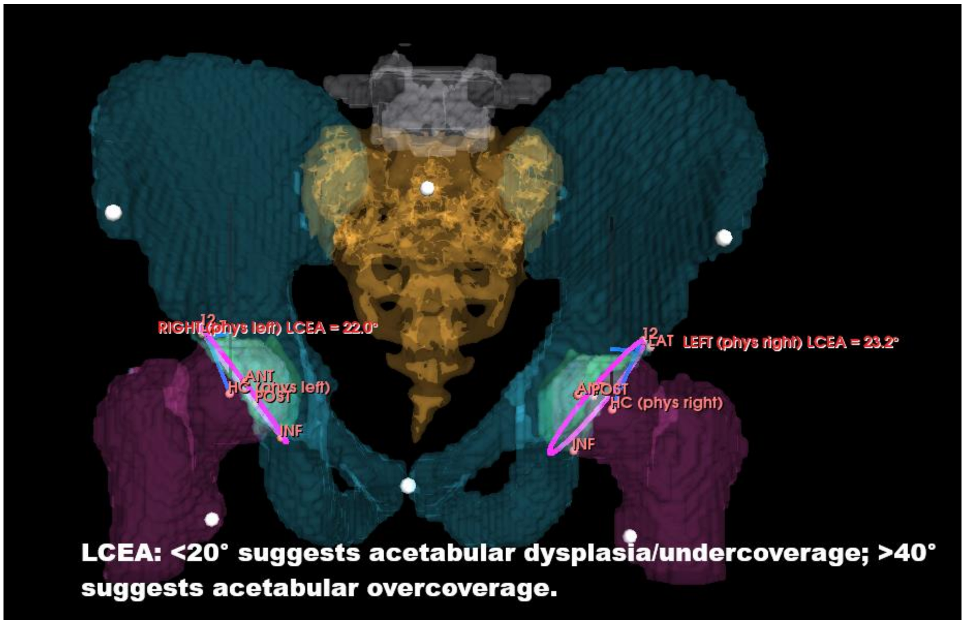


**Figure 2. Segmentation-derived lateral center-edge angle measurement.** LCEA is calculated from the segmented acetabular rim and femoral-head center in a fixed AP/coronal plane.

**The acetabular index**, (sharp angel) , quantifies acetabular roof inclination and may also differ between radiographs and CT in dysplasia assessment [5]. The classical acetabular index/Tönnis angle remains rooted in established dysplasia measurement principles [6]. In the present workflow, the closest acetabular patch was used to extract the rim, identify the most lateral and medial-inferior rim points, and calculate the acute roof angle in the standardized camera plane (Figure 3).

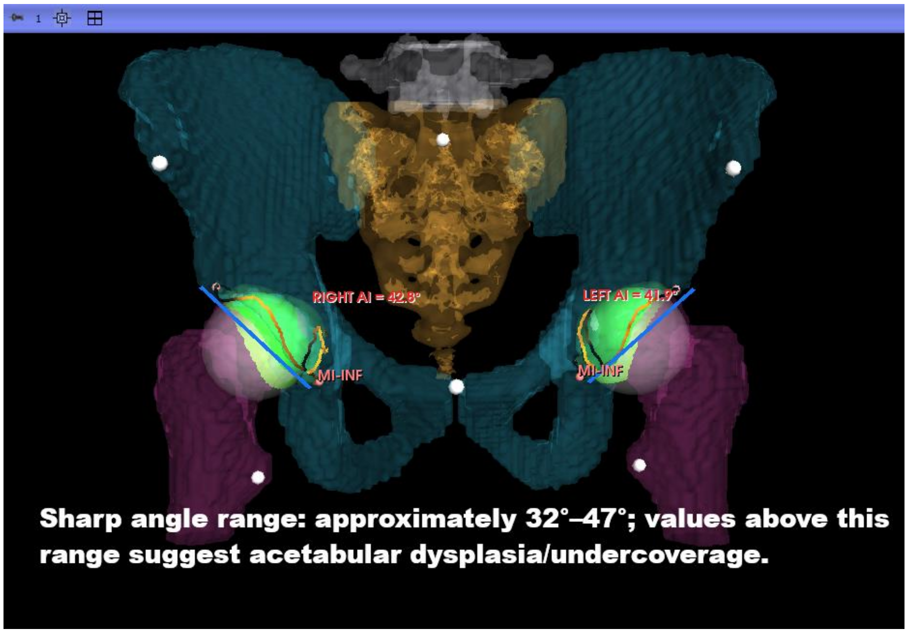


**Figure 3. Segmentation-derived acetabular index measurement.** The acetabular index is calculated from the medial-inferior and lateral acetabular rim points in the standardized camera plane.

**Acetabular version** describes the axial opening orientation of the socket according to established definitions of acetabular orientation [7]. In the present workflow, version was estimated on the femoral-head axial level using segmented anterior-wall, posterior-wall, and contralateral posterior-wall references, with the angle calculated from same-side anterior–posterior wall geometry relative to the transverse posterior reference (Figure 4).

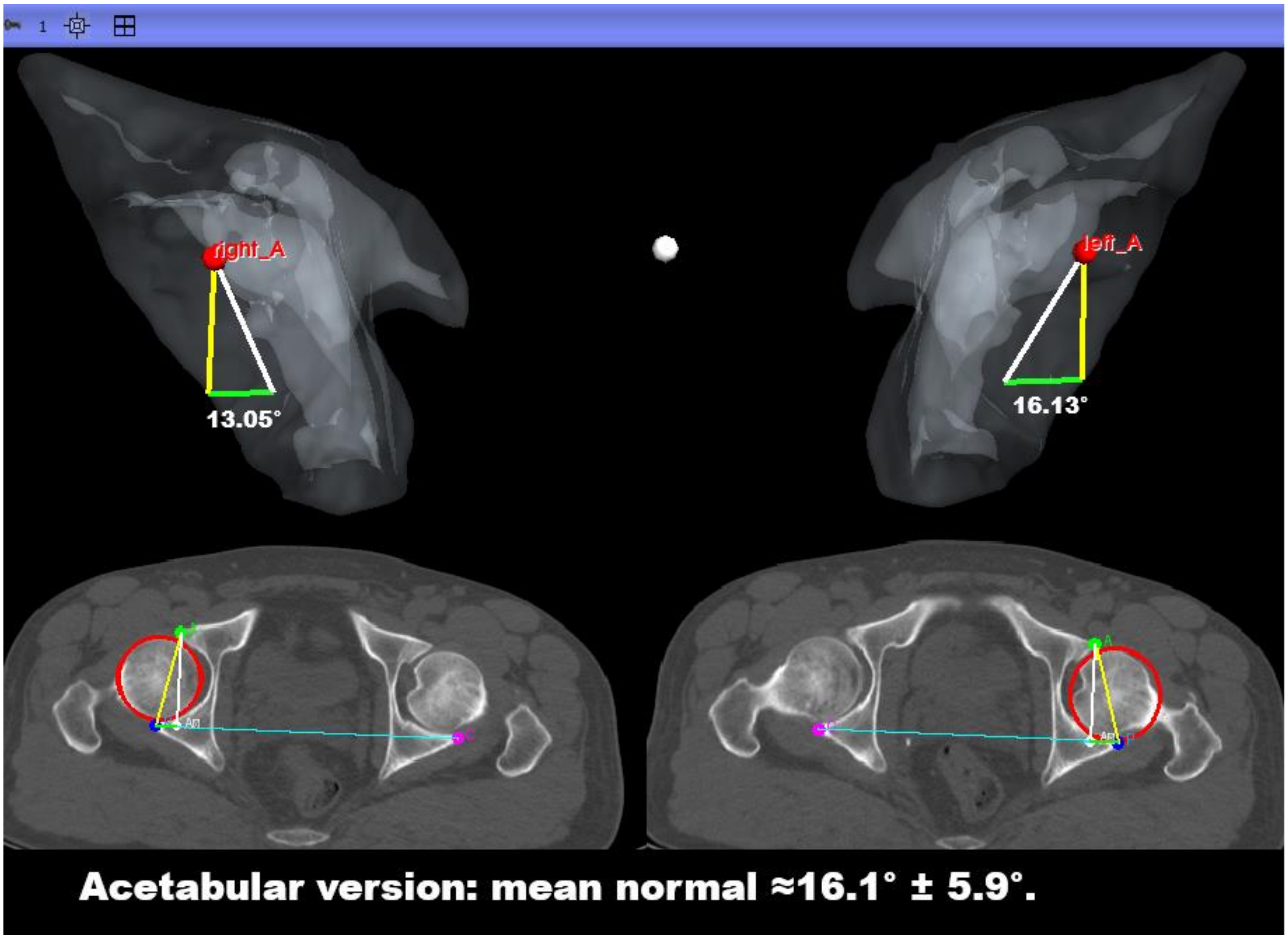


**Figure 4. Segmentation-derived acetabular version measurement.** Acetabular version is estimated from segmented anterior- and posterior-wall references at the femoral-head axial level.

Clinically, this combined coverage package may refine phenotyping of dysplasia, pincer morphology, anterior–posterior wall imbalance, and mixed femoroacetabular deformity.

Its novelty is not 3D acetabular coverage analysis itself, but the automated derivation of regional coverage maps from a validated segmentation tree, reducing manual boundary tracing and linking coverage percentages, clockface localization, LCEA, roof inclination, and acetabular version in one post-segmentation workflow.

#### Proximal femoral and reconstruction-related metrics

Proximal femoral and pelvic metrics were included because hip mechanics and reconstruction depend on coronal alignment, torsion, offset, limb-length symmetry, and pelvic posture.

- **Neck–shaft angle ( NSA )**

The neck–shaft angle quantifies the coronal relationship between the femoral neck and shaft and is used to describe proximal femoral morphology, including coxa vara and coxa valga [8]. In the present workflow, NSA was calculated from the segmented femoral head–neck axis and an adaptive femoral shaft axis selected from the available proximal or mid-shaft region (Figure 5). This output supports proximal femoral morphology assessment, bilateral comparison, osteotomy planning, and implant geometry evaluation.

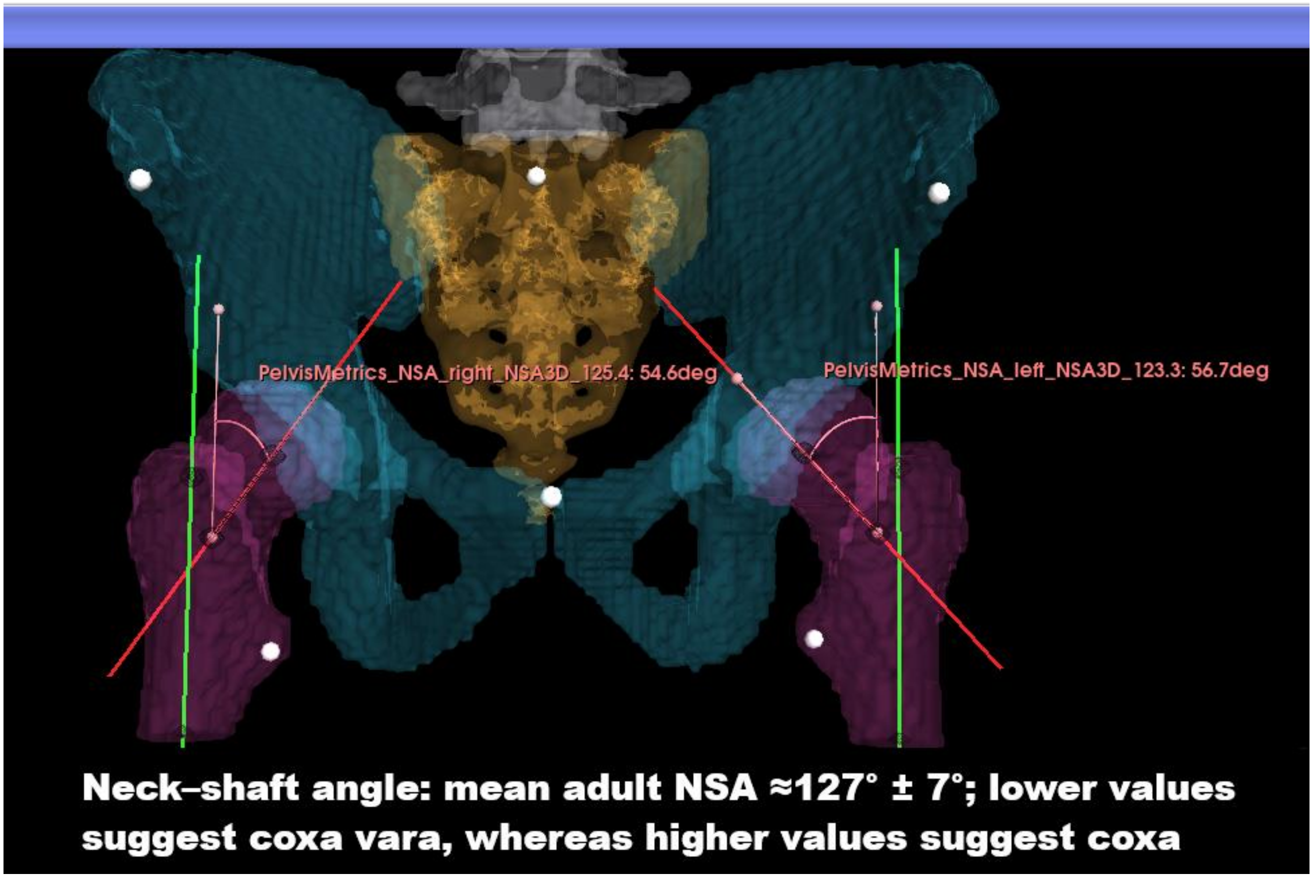


**Figure 5. Segmentation-derived neck-shaft angle measurement.** NSA is calculated from the segmented femoral head-neck axis and adaptive femoral shaft axis.

- **Femoral offset**

Femoral offset is the perpendicular distance from the femoral-head center to the femoral shaft axis and is important for hip biomechanics, abductor function, gait, stability, and THA reconstruction [9,10].

In the present workflow, femoral offset was measured by projecting the segmented femoral-head center and adaptive shaft axis into the fixed frontal plane and calculating the perpendicular distance (Figure 6). This output supports templating, reconstruction assessment, postoperative comparison, and side-to-side symmetry evaluation.

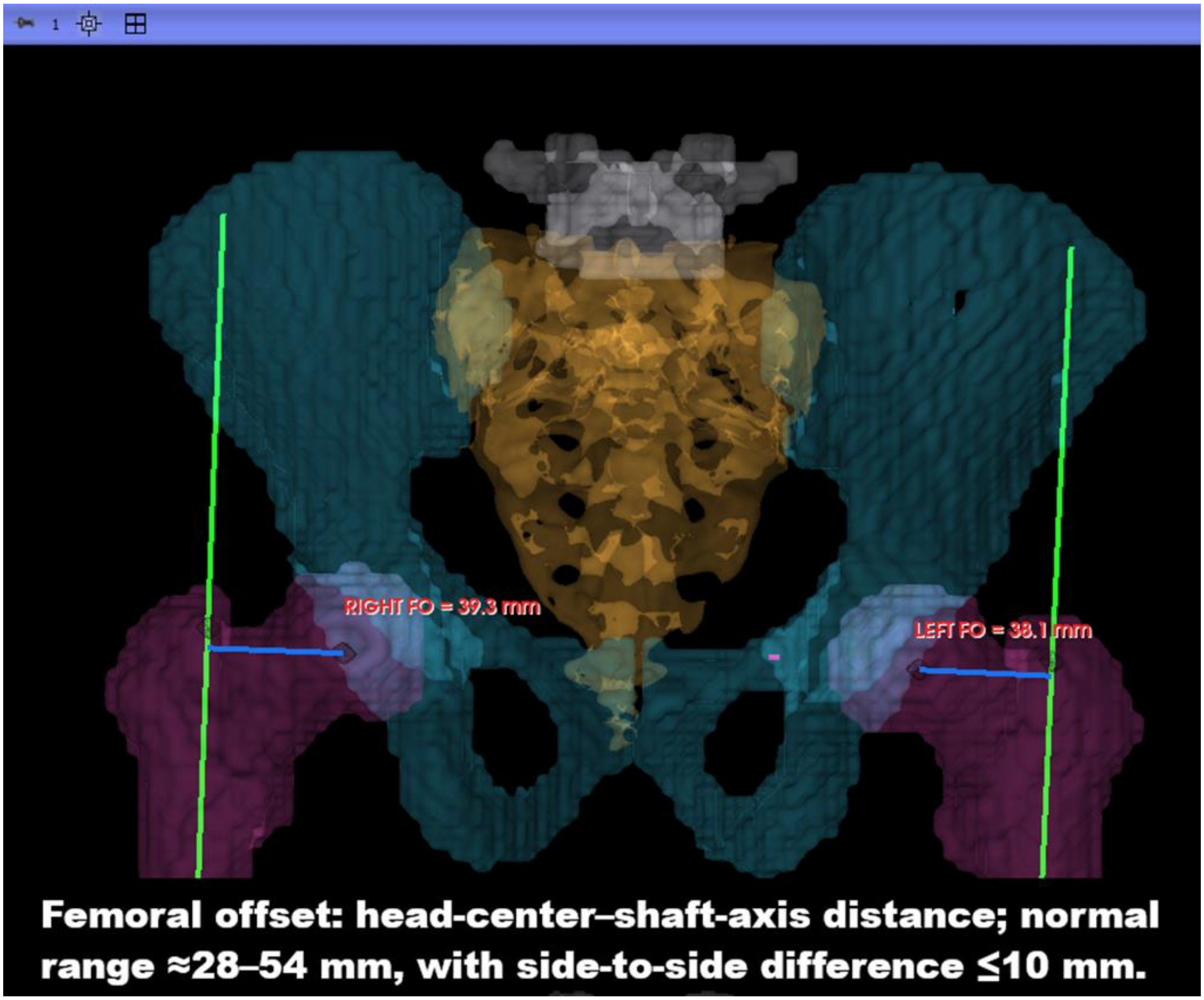


**Figure 6. Segmentation-derived femoral offset measurement.** Femoral offset is measured as the perpendicular distance from the femoral-head center to the adaptive femoral shaft axis.

- **Limb-length discrepancy**

Limb-length discrepancy reflects side-to-side length inequality and is clinically relevant because residual discrepancy affects gait, patient satisfaction, function, and perceived reconstruction quality. [11,10]

In the present workflow, LLD was estimated by measuring the 3D ASIS-to-lesser-trochanter distance on each side and reporting the left–right difference.

This output provides a rapid internal bilateral comparison for preoperative, postoperative, and reconstruction-related assessment (Figure 7).

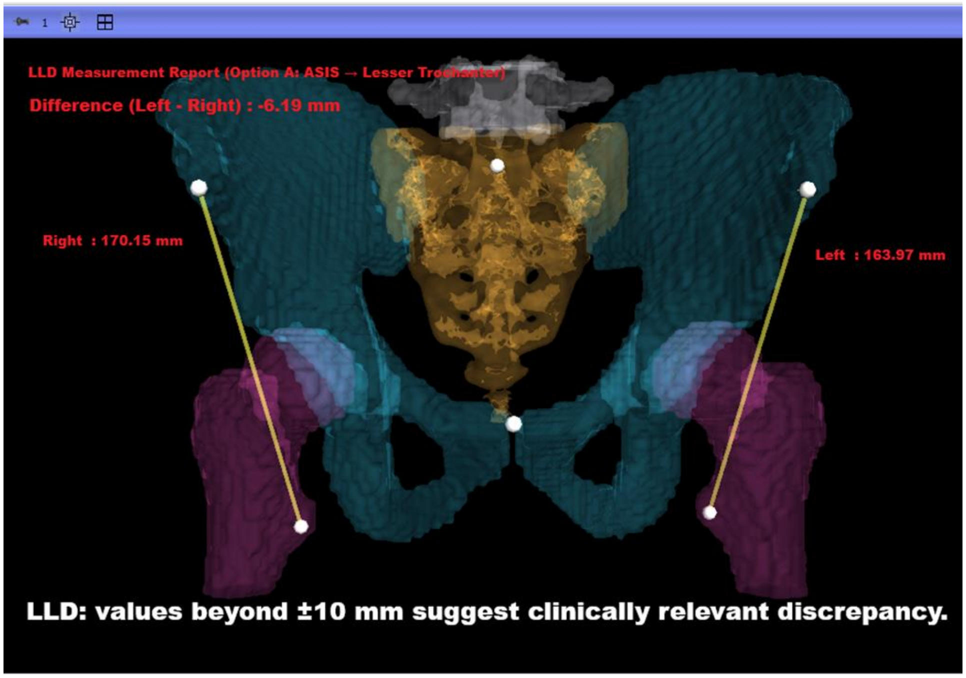


**Figure 7. Segmentation-derived limb-length discrepancy measurement.** LLD is estimated from bilateral ASIS-to-lesser-trochanter distances and reported as the side-to-side difference.

#### Pelvic tilt

Pelvic tilt describes sagittal pelvic orientation and is clinically relevant because pelvic posture changes functional acetabular orientation and cup-position interpretation [12,13]. In the present workflow, pelvic tilt was calculated from the anterior pelvic plane defined by the segmented ASIS landmarks and symphysis pubis (Figure 8).

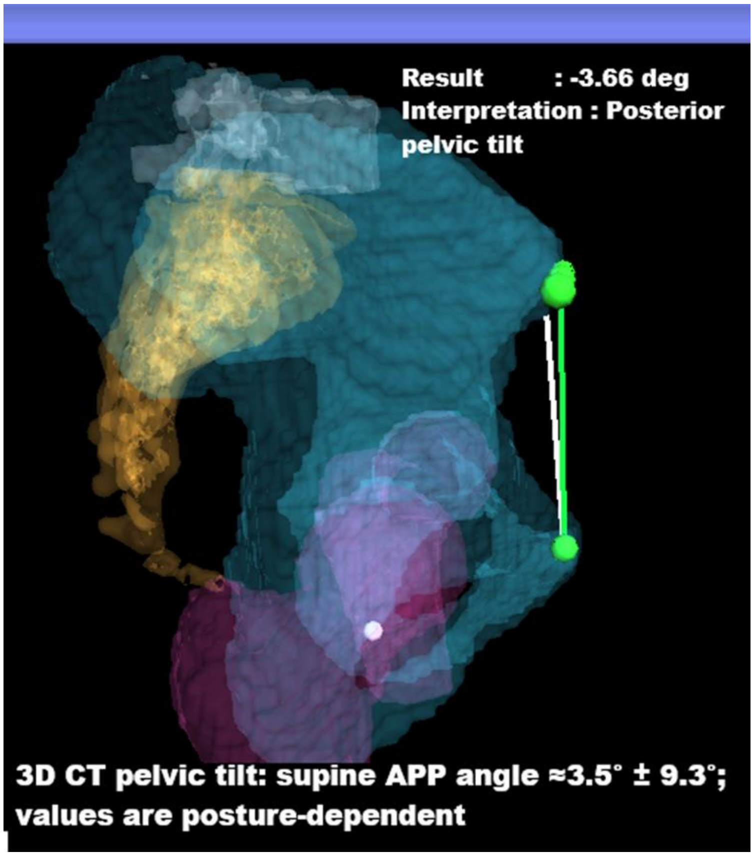


**Figure 8. Segmentation-derived pelvic tilt and reconstruction-related metrics.** Pelvic tilt is calculated from the anterior pelvic plane defined by the ASIS landmarks and symphysis pubis.

#### Pelvic bone-quality assessment

Pelvic bone-quality assessment supports osteoporotic-fracture risk stratification and surgical planning, as diminished bone density and regional bone stock are linked to implant purchase, screw pull-out, loosening, reduction collapse, and fixation failure [14,15,16]. Opportunistic CT screening with vertebral HU correlates with osteoporosis, but it does not describe the heterogeneous pelvic bone stock used for fixation or arthroplasty [14]. Pelvic cancellous HU varies regionally, with denser load-transfer areas such as the acetabular roof and sciatic buttress differing from the pubis and ischium [17].

In the present workflow, PBQI sampled the segmented weight-bearing chain: L5 body, sacrum, bilateral sciatic buttress, acetabular roof, femoral head, and calcar. Each ROI was extracted from the finalized segmentation tree, internally filled, eroded toward trabecular bone, and analyzed by voxel HU distribution. HU values were summarized as mean HU and four density bands: <150, 150–250, 250–350, and >350 HU.

The immediate output is a regional heat-map and table showing local bone stock and side-to-side asymmetry.

This framework could evolve into a composite pelvic bone-quality index combining mean HU, HU-band fractions, regional asymmetry, and load-bearing ROI patterns. [18,19]

The novelty is shifting from single-level HU or DXA screening to a segmentation-guided, multi-region pelvic index aligned with surgical load-bearing anatomy.

PBQI remains a feasibility framework and must be validated against DXA/QCT, fracture phenotype, screw purchase, fixation failure, implant loosening, biomechanical testing, and clinical outcomes before clinical use (Figure 9).

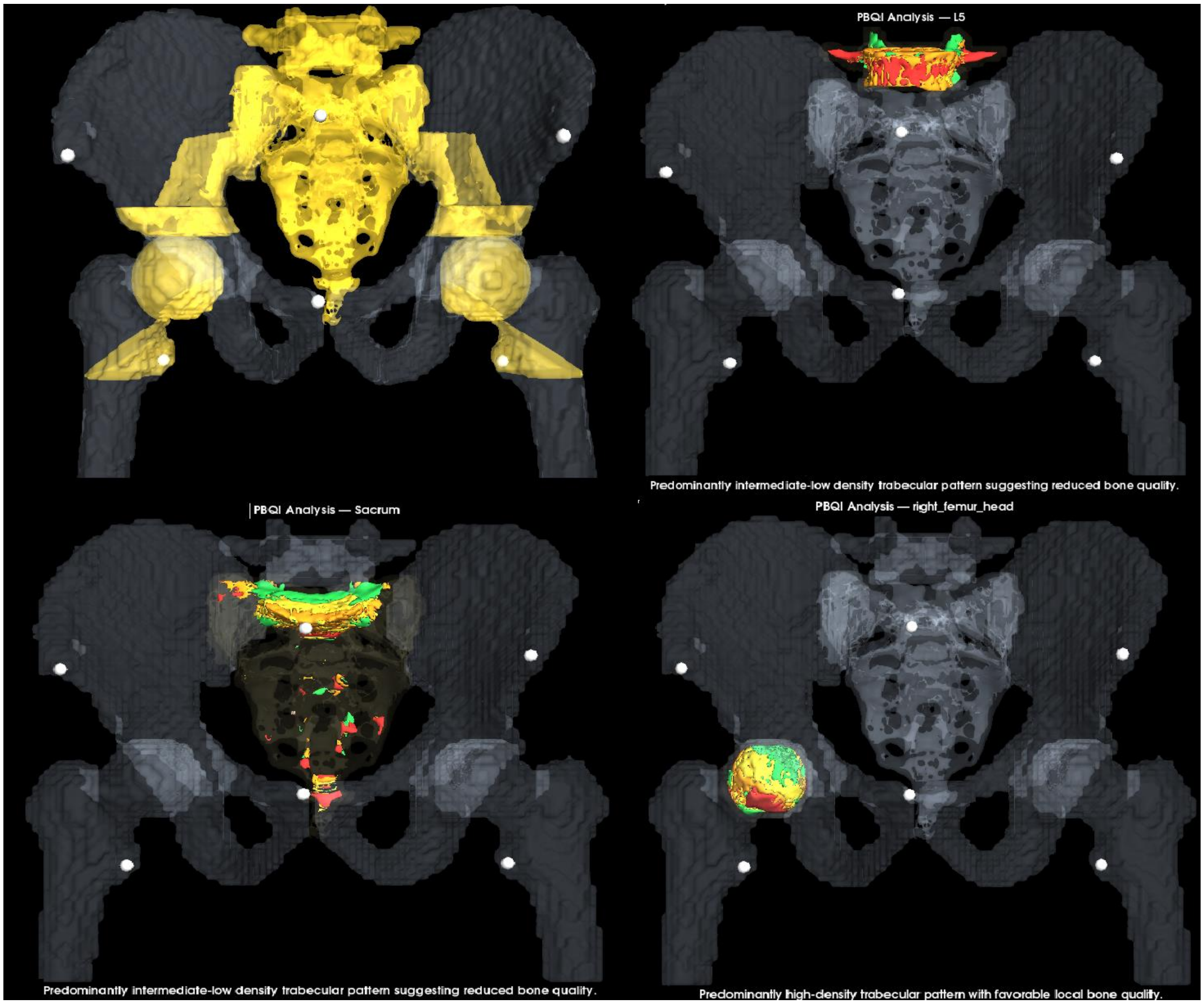


**Figure 9. Pelvic bone-quality assessment across segmented load-bearing regions.** Four HU bands summarize local pelvic bone stock along the segmented load-bearing chain from L5 to calcar.

#### Arthroplasty-planning application

CT-based 3D THA planning improves anatomical understanding, implant-size prediction, component positioning, and reconstruction strategy [20]. However, conventional CT-based 3D planning may require manual segmentation, repeated component adjustment, operator experience, and substantially longer templating time; Huo et al. reported mean planning time of 32.07 ± 2.41 min for 3D Mimics versus 3.91 ± 0.64 min for AI-assisted 3D planning. [21]

In the present workflow, arthroplasty planning represents a direct surgical extension of pelvic CT subsegmentation.The module established an APP-based pelvic coordinate system from segmented pelvic landmarks and used this frame for standardized planning. The cup-planning tool derived the acetabular center, press-fit cup size, inclination, version, rim engagement, and peri-acetabular HU from the segmented acetabular contact region. The femoral-planning tool used STL implant components and segmented femoral references to align the stem, fit it to the medullary isthmus, and report head size, stem size, femoral version, combined anteversion, and proximal bone morphology. The workflow generated implant-in-situ visualization, LLD/offset summaries, bone-quality context, femoral-fit outputs, and a consolidated planning report (Figure 10).

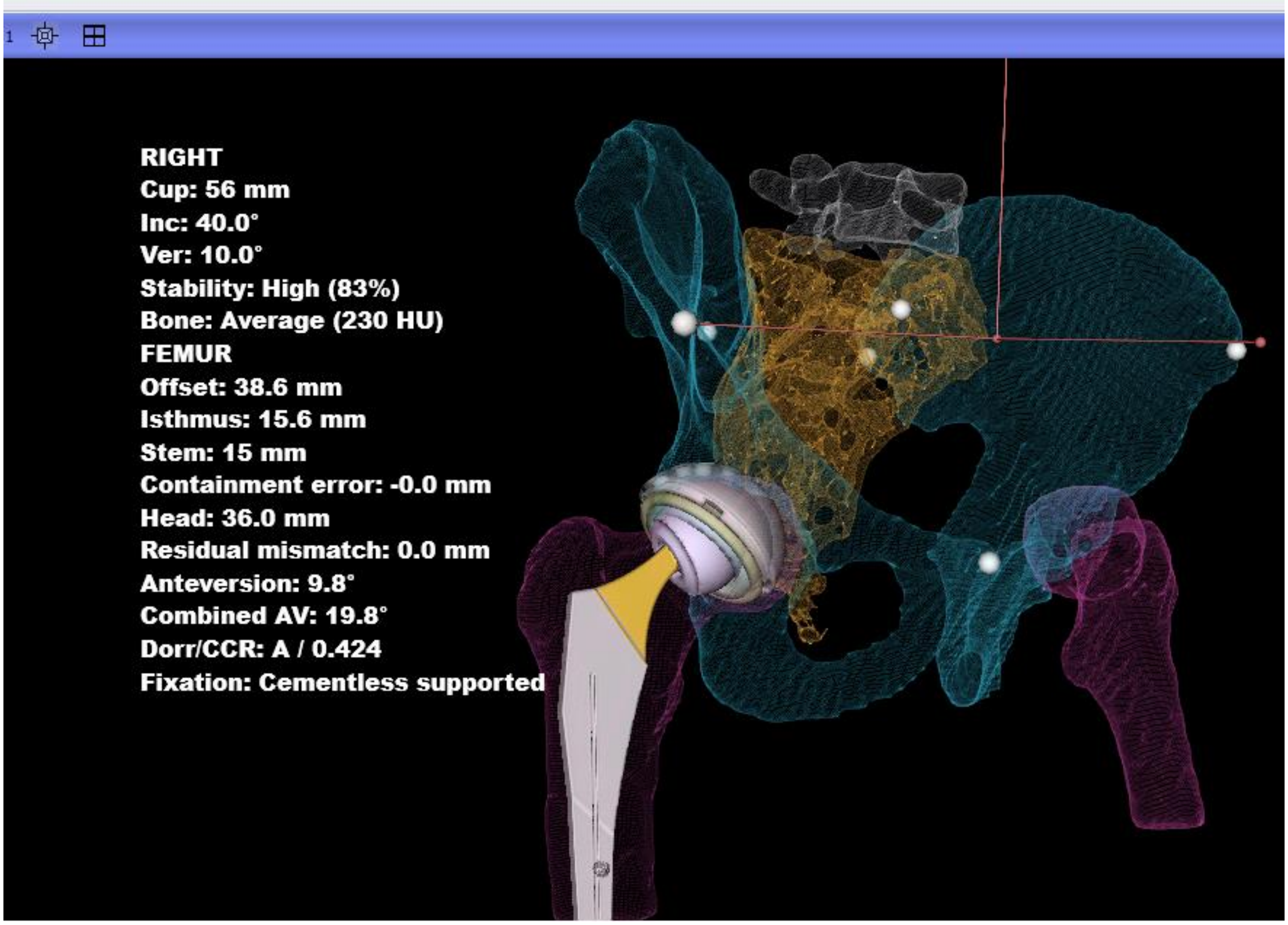


**Figure 10. Arthroplasty-planning workflow derived from pelvic CT subsegmentation.** The same segmented anatomy drives pelvic-frame standardization, cup planning, femoral stem fitting, implant visualization, and final reporting.

The novelty is a one-click segmentation-derived THA planning workflow that integrates pelvic orientation, cup planning, femoral stem fitting, HU context, implant visualization, and reporting, aiming to reduce manual CT-planning burden and improve planning consistency and accuracy.

These outputs demonstrate feasibility only; implant-selection rules, stability metrics, HU thresholds, and final reconstruction parameters require validation against surgeon planning, navigation or robotic systems, postoperative imaging, and clinical outcomes.

#### CT-based screw-corridor planning

CT-based screw-corridor planning is essential for defining safe intraosseous trajectories in pelvic and acetabular fixation, where narrow osseous pathways leave little tolerance for cortical breach, joint penetration, or neurovascular injury [22,23,24,25].

In the present workflow, the finalized Pelvis Segmentation Tree was used to define screw entry points, target regions, corridor axes, diameters, lengths, and regional HU context for preoperative corridor screening, implant sizing, entry localization, and bone-quality assessment (Figure 11).

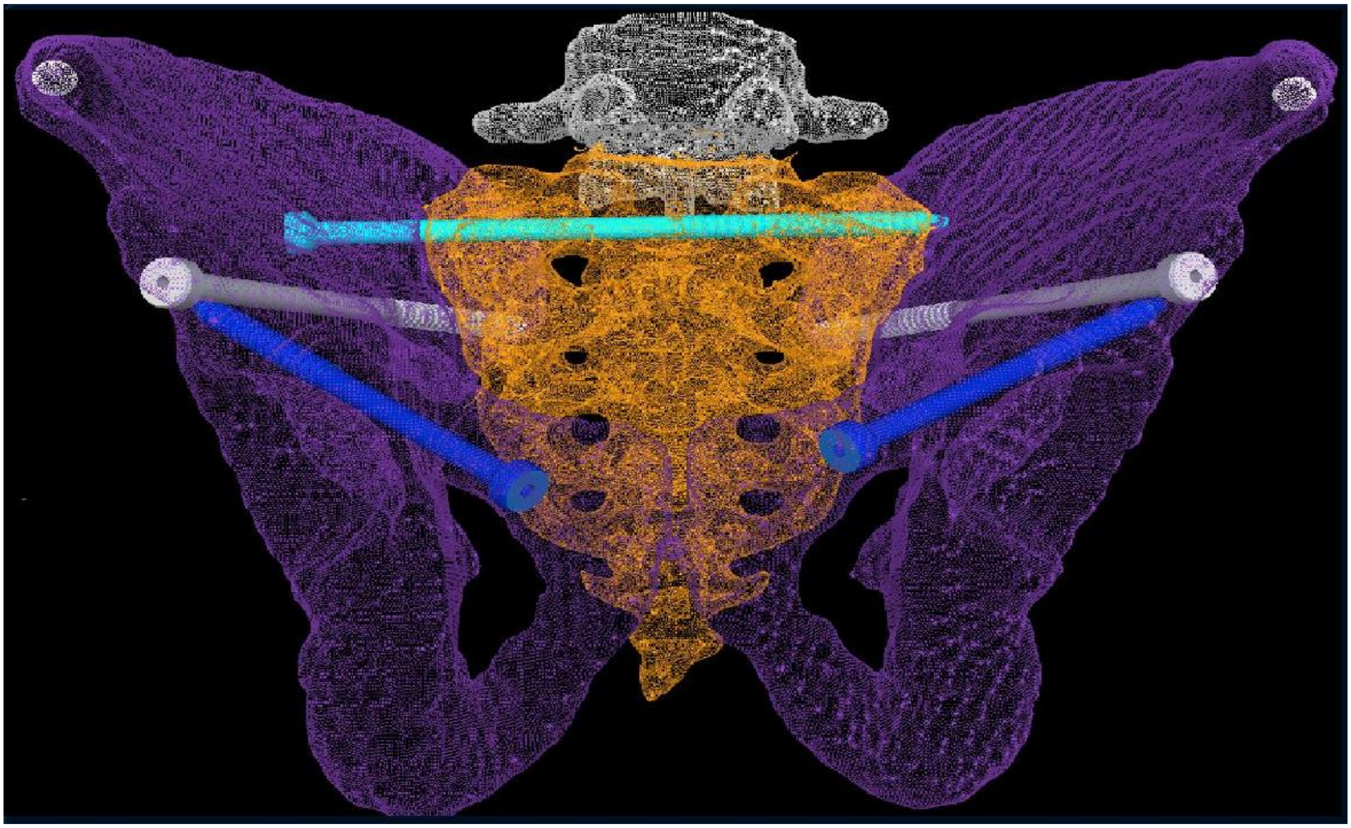

**Figure 11. CT-based screw-corridor planning from segmented pelvic anatomy.** Segmented entry regions, target zones, corridor axes, diameters, lengths, and local HU context support preoperative screw-corridor screening.

#### Deep-learning dataset expansion

Recent pelvic segmentation studies highlight the need for robust CT labels to train and test AI models across normal anatomy, anatomical variation, and fracture morphology [26,27].

In the present workflow, validated deterministic subsegmentation can generate large-scale initial masks for normal pelvic CTs. Abnormal and fracture cases can then be semi-automatically segmented and manually refined to create pathology-aware labels.

The next step is extension to the main pelvic anatomical structures and clinically relevant subregions, including the acetabulum, ilium, sacrum, pubis, anterior column, posterior column, and additional subsegmentations.

These curated labels could support automated pelvic segmentation, abnormality detection, fracture localization, displacement measurement, bone-loss assessment, and pelvic or acetabular fracture classification.

### References


1. DeFroda SF, Alter TD, Lambers F, Malloy P, Clapp IM, Chahla J, et al. Quantification of acetabular coverage on 3-dimensional reconstructed CT scan bone models in patients with femoroacetabular impingement syndrome: a descriptive study. Orthop J Sports Med. 2021;9(11):23259671211049457. doi:10.1177/23259671211049457.

2. Kamenaga T, Ritacco L, Slullitel PA, Nahal C, Nepple JJ, Clohisy JC, et al. Prediction of 3-dimensional coverage surface area of the femoral head in hip dysplasia through conventional computed tomography. Orthop J Sports Med. 2024;12(3):23259671241234684. doi:10.1177/23259671241234684.

3. Upasani VV, Bomar JD, Bandaralage H, Doan JD, Farnsworth CL. Assessment of three-dimensional acetabular coverage angles. J Hip Preserv Surg. 2020;7(2):305-312. doi:10.1093/jhps/hnaa026.

4. Nerys-Figueroa J, Kahana-Rojkind AH, Parsa A, Maldonado D, Quesada-Jimenez R, Domb BG. The measurement of the lateral center-edge angle is underestimated on radiographs compared with 3-dimensional CT. Arthrosc Sports Med Rehabil. 2025;7(1):101005. doi:10.1016/j.asmr.2024.101005.

5. Nerys-Figueroa J, Kahana-Rojkind AH, Parsa A, Walsh EG, Lambers F, Domb BG. Radiographs underestimate lateral center-edge angle and Tönnis angle measurements compared to computed tomography scan in assessment of borderline and frank acetabular dysplasia. Arthroscopy. 2025;41(7):2343-2350. doi:10.1016/j.arthro.2024.10.038.

6. Tönnis D. Congenital Dysplasia and Dislocation of the Hip in Children and Adults. Berlin, Heidelberg: Springer-Verlag; 1987.

7. Murray DW. The definition and measurement of acetabular orientation. J Bone Joint Surg Br. 1993;75(2):228-232. doi:10.1302/0301-620X.75B2.8444942.

8. Fischer CS, Kühn JP, Völzke H, Ittermann T, Gümbel D, Kasch R, et al. The neck–shaft angle: an update on reference values and associated factors. Acta Orthop. 2020;91(1):53-57. doi:10.1080/17453674.2019.1690873.

9. Oommen AT. Offset restoration in total hip arthroplasty: Important: A current review. World J Orthop. 2024;15(8):696-703. doi:10.5312/wjo.v15.i8.696.

10. Enke O, Levy YD, Bruce WJM. Accuracy of leg length and femoral offset restoration after total hip arthroplasty with the utilisation of an intraoperative calibration gauge. Hip Int. 2020;30(3):296-302. doi:10.1177/1120700019836383.

11. Bakr H, Mahran M. Assessment of restoration of leg length and femoral offset after total hip arthroplasty. Egypt Orthop J. 2021;56(3):148-152. doi:10.4103/eoj.eoj_91_21.

12. Lembeck B, Müller O, Reize P, Wülker N. Pelvic tilt makes acetabular cup navigation inaccurate. Acta Orthop. 2005;76(4):517-523. doi:10.1080/17453670510041501.

13. Stadnyk M, Liu T, Fallahi Arezodar F, Westover L, Carvajal Alba JA, Masson E, et al. Analysis of four methods of measuring three-dimensional pelvic tilt in the lateral decubitus position. Med Biol Eng Comput. 2020;58(10):2387-2396. doi:10.1007/s11517-020-02235-4.

14. Pickhardt PJ, Pooler BD, Lauder T, del Rio AM, Bruce RJ, Binkley N. Opportunistic screening for osteoporosis using abdominal computed tomography scans obtained for other indications. Ann Intern Med. 2013;158(8):588-595. doi:10.7326/0003-4819-158-8-201304160-00003.

15. Rommens PM, Wagner D, Hofmann A. Stabilization of osteoporotic pelvis and acetabular fractures. Indian J Orthop. 2025;59(3):375-388. doi:10.1007/s43465-024-01315-z.

16. Li J, Zhang Z, Xie T, Song Z, Song Y, Zeng J. The preoperative Hounsfield unit value at the position of the future screw insertion is a better predictor of screw loosening than other methods. Eur Radiol. 2023;33(3):1526-1536. doi:10.1007/s00330-022-09157-9.

17. Inagaki N, Tanaka T, Udaka J, Akiyama S, Matsuoka T, Saito M. Distribution of Hounsfield unit values in the pelvic bones: a comparison between young men and women with traumatic fractures and older men and women with fragility fractures. BMC Musculoskelet Disord. 2022;23:305. doi:10.1186/s12891-022-05263-3.

18. Chaisen M, Sritara C, Chitrapazt N, Suppasilp C, Chamroonrat W, Promma S, et al. Opportunistic screening for osteoporosis by CT as compared with DXA. Diagnostics (Basel). 2024;14(24):2846. doi:10.3390/diagnostics14242846.

19. Yang T-J, Wen P-P, Ye X, Wu X-F, Zhang C, Sun S-Y, et al. CT Hounsfield units in assessing bone and soft tissue quality in the proximal femur: a systematic review focusing on osteonecrosis and total hip arthroplasty. PLoS One. 2025;20(3):e0319907. doi:10.1371/journal.pone.0319907.

20. Moralidou M, Di Laura A, Henckel J, Hothi H, Hart AJ. Three-dimensional pre-operative planning of primary hip arthroplasty: a systematic literature review. EFORT Open Rev. 2020;5(12):845-855. doi:10.1302/2058-5241.5.200046.

21. Huo J, Huang G, Han D, Wang X, Bu Y, Chen Y, et al. Value of 3D preoperative planning for primary total hip arthroplasty based on artificial intelligence technology. J Orthop Surg Res. 2021;16:156. doi:10.1186/s13018-021-02294-9.

22. Noser H, Radetzki F, Stock K, Mendel T. A method for computing general sacroiliac screw corridors based on CT scans of the pelvis. J Digit Imaging. 2011;24(4):665-671. doi:10.1007/s10278-010-9327-0.

23. Mendel T, Noser H, Wohlrab D, Stock K, Radetzki F. CT-based 3-D visualisation of secure bone corridors and optimal trajectories for sacroiliac screws. Injury. 2013;44(7):957-963. doi:10.1016/j.injury.2012.11.013.

24. Lu Q, Zhou R, Gao S, Liang A, Yang M, Yang H. CT-scan based anatomical study as a guidance for infra-acetabular screw placement. BMC Musculoskelet Disord. 2021;22:478. doi:10.1186/s12891-021-04419-x.

25. Yu K, Zhou R, Gao S, Liang A, Yang M, Yang H. The placement of percutaneous retrograde acetabular posterior column screw based on imaging anatomical study of acetabular posterior column corridor. J Orthop Surg Res. 2022;17:492. doi:10.1186/s13018-022-03347-3.

26. Liu P, Han H, Du Y, Zhu H, Li Y, Gu F, et al. Deep learning to segment pelvic bones: large-scale CT datasets and baseline models. Int J Comput Assist Radiol Surg. 2021;16(5):749-756. doi:10.1007/s11548-021-02363-8.

27. Lee JM, Park JY, Kim YJ, et al. Deep-learning-based pelvic automatic segmentation in pelvic fractures. Sci Rep. 2024;14:12258. doi:10.1038/s41598-024-63093-w.
